# Dynamics of the blood plasma proteome during hyperacute HIV-1 infection

**DOI:** 10.1101/2024.04.30.24306610

**Authors:** Jamirah Nazziwa, Eva Freyhult, Mun-Gwan Hong, Emil Johansson, Filip Årman, Jonathan Hare, Kamini Gounder, Melinda Rezeli, Tirthankar Mohanty, Sven Kjellström, Anatoli Kamali, Etienne Karita, William Kilembe, Matt A Price, Pontiano Kaleebu, Susan Allen, Eric Hunter, Thumbi Ndung’u, Jill Gilmour, Sarah L. Rowland-Jones, Eduard Sanders, Amin S. Hassan, Joakim Esbjörnsson

**Affiliations:** Department of Translational Medicine, Lund University, Sweden; Lund University Virus Centre, Lund University, Sweden; Department of Cell and Molecular Biology, National Bioinformatics Infrastructure Sweden, Science for Life Laboratory, Uppsala University, Uppsala, Sweden; National Bioinformatics Infrastructure Sweden, Science for Life Laboratory, Department of Biochemistry and Biophysics, Stockholm University, Stockholm, Sweden; BioMS−Swedish National Infrastructure for Biological Mass Spectrometry, Lund University, Lund, Sweden; IAVI Human Immunology Laboratory, Imperial College, London, United Kingdom; IAVI, New York, New York, USA, and Nairobi, Kenya; Africa Health Research Institute, Durban, South Africa; HIV Pathogenesis Programme, The Doris Duke Medical Research Institute, University of KwaZulu-Natal, Durban, South Africa; Division of Infection Medicine, Department of Clinical Sciences Lund, Faculty of Medicine, Lund University, Lund, Sweden; Rwanda/Zambia HIV Research Group, Kigali, Rwanda, and Lusaka, Zambia; UCSF Department of Epidemiology and Biostatistics, San Francisco, California, USA; Medical Research Council/Uganda Virus Research Institute, Uganda, and London School of Hygiene and Tropical Medicine, London, United Kingdom; Emory Vaccine Center, Emory University, Atlanta, Georgia, USA; Ragon Institute of Massachusetts General Hospital, Massachusetts Institute of Technology and Harvard University, Cambridge, Massachusetts, USA; Division of Infection and Immunity, University College London, London, United Kingdom; Nuffield Department of Clinical Medicine, University of Oxford, UK; KEMRI/Wellcome Trust Research Programme, Kilifi, Kenya; The Aurum Institute, Johannesburg, South Africa

## Abstract

HIV-1 remains incurable and there is no effective vaccine towards the infection. A main challenge for this is the lack of a holistic understanding of the myriad of complex virus-host interactions during hyperacute HIV-1 infection (hAHI), and how these contribute to tissue damage and pathogenesis. Here, 1293 blood plasma proteins were quantified from 157 linked plasma samples collected before, during, and after hAHI of 54 volunteers from four sub-Saharan African countries. Six distinct longitudinal expression profiles were identified, of which four demonstrated a consistent decrease in protein levels following HIV-1 infection. Differentially expressed proteins were involved in inflammation, innate immunity, cell motility, and actin cytoskeleton reorganisation. Specifically, decreased levels of Zyxin, Secretoglobin family 1A member 1, and Pro-platelet basic protein were associated with acute retroviral syndrome; Rho GTPase activating protein 18, Annexin A1, and Lipopolysaccharide binding protein with viral load; and Hepsin, Protein kinase C beta, and Integrin subunit beta 3 with disease progression. This is the first holistic characterisation of within-patient blood plasma proteome dynamics during the first weeks of HIV-1 infection and presents multiple potential blood biomarkers and targets for prophylactic and therapeutic HIV-1 interventions.

## INTRODUCTION

While the blood plasma proteome typically remains stable in healthy individuals, perturbations have been documented in response to different infections, including severe acute respiratory syndrome coronavirus 2, and bacterial and viral pneumonia^1,2^. Understanding differential expression of plasma proteomics in these infections played a pivotal role in informing diagnostic, prophylactic, and therapeutic interventions^3,4^. In HIV-1 infection, virus-host interactions during the earliest stages of infection – the hyperacute HIV-1 infection (hAHI, defined as the period from onset of plasma viremia to peak viral load) – triggers a complex network of cell and tissue signalling that manifests as rapid systemic immune activation and reorganisation of cellular microenvironments^5–8^. A part of this has been associated with a cytokine storm, and some infected individuals show symptoms of acute retroviral syndrome (ARS)^9–12^. These events contribute to a significant loss of CD4+ T-cells and germinal centres and play a critical role in shaping HIV-1 disease pathogenesis^6,8,13–15^. Indeed, disease progression to AIDS in HIV-1 infected individuals varies widely, ranging from a few months to several decades, and events during hAHI have been suggested to strongly influence the rate of disease progression^16–18^. In blood, HIV-1 viraemia is detectable approximately a week after infection, reaching a peak viral load (VL) of millions of virus particles per ml 3-4 weeks after infection^5,6^. During this period, a reservoir of latently infected cells is also established, which remains one the main obstacles of curing HIV-1^19^. After the peak VL, the viraemia gradually decreases and plateaus at a set-point level ∼30-65 days after infection^5,6^.

While pro-inflammatory and antiviral cytokines and chemokines have been studied extensively, longitudinal studies investigating blood plasma proteins during hAHI are lacking^13,20,21^. Recent advancements in data-independent acquisition mass spectrometry (DIA-MS) based proteomics has enabled simultaneous identification and quantification of thousands of proteins in plasma across a large dynamic range, significantly expanding the potential of detectable biomarkers^22^. Furthermore, the cellular and tissue expression patterns of these proteins have been mapped in a human protein atlas, facilitating analysis of proteome dynamics in response to infections^23^. In this study, data and samples collected from two well-characterised HIV-1 incidence cohorts from four sub-Sahara African countries were analysed to elucidate dynamics of blood plasma proteome before, during and after hAHI, and to determine associations with ARS, viral load responses, and HIV-1 disease progression.

## RESULTS

### Study participants

Overall, 54 participants from the East African (International AIDS Vaccine Initiative, IAVI, cohort, n=39), and South African (Females Rising through Education, Support, and Health, FRESH, and HIV Pathogenesis Programme, HPP, cohorts, Durban, n=15) met the eligibility criteria, and were included in the analyses (Fig. 1)^24–27^. Participants contributed 157 longitudinally linked plasma samples from three time points including Visit 0 (V0), collected at a median of 62 days prior to HIV-1 infection (interquartile range [IQR] 28-106 days); V1, median 10 days after HIV-1 infection (IQR 10-14); and V2, median 31 days after HIV-1 infection (IQR 28-37)^28^. Most participants were male (n=34, 63%), aged below 25 years (n=32, 59%), from Kenya (n=32, 59%), infected with HIV-1 sub-subtype A1 (n=31, 57%), and identified as men who have sex with men (MSM, n=28, 52%, Extended Data Table 1).

**Fig. 1.**
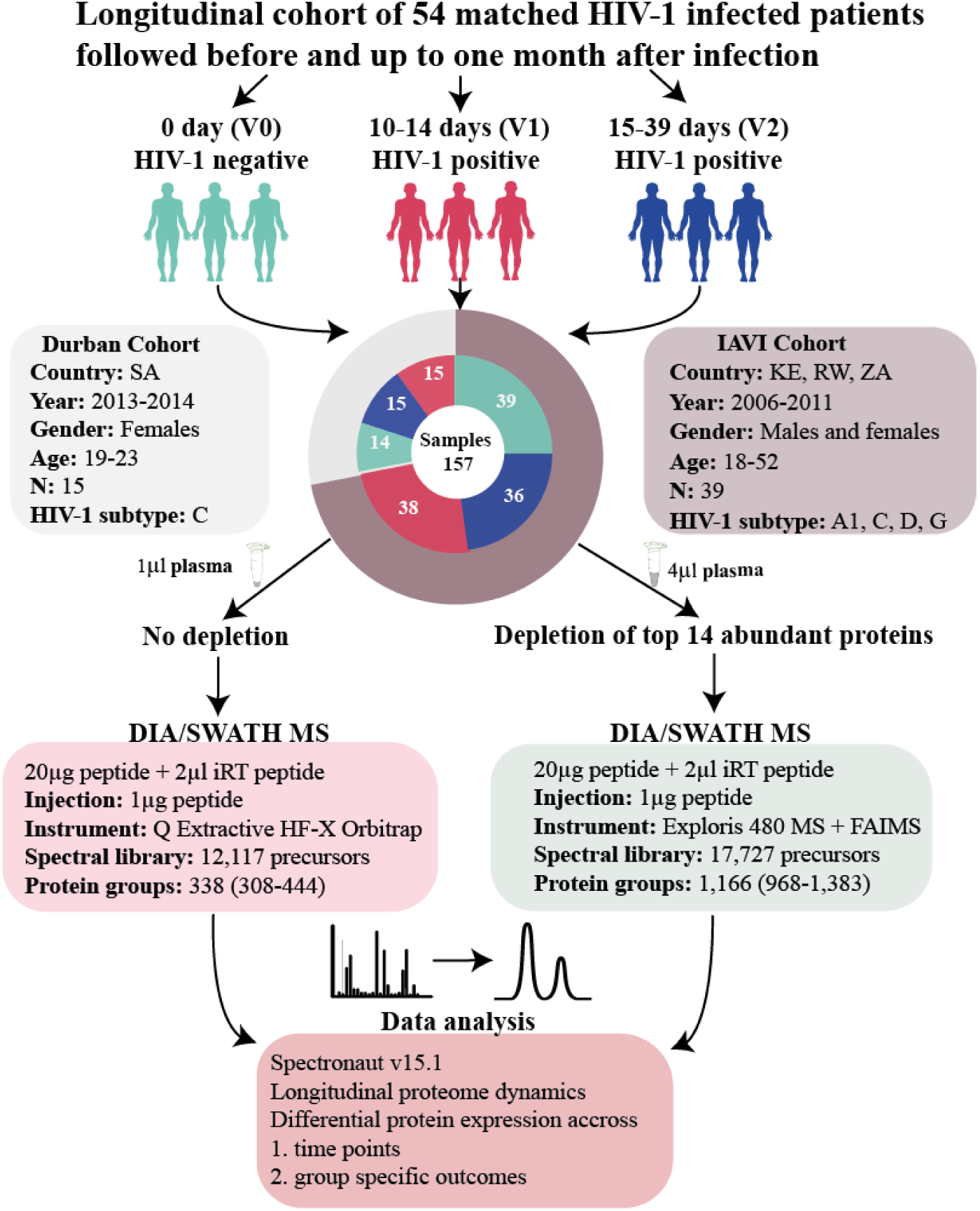
Characteristics of the study participants. The flowchart illustrates the longitudinal sampling approach adopted in the study and outlines the workflow for processing proteomic data. A total of 54 individuals from two geographical regions contributed three matched each. These samples underwent plasma preparation with and without depletion of top 14 abundant proteins, followed by DIA/SWATH LC-MS/MS analysis, and subsequent computational analyses. Arrows denote the flow of samples and processing steps. Abbreviations: SA, South Africa; KE, Kenya; DIA, data-independent acquisition; SWATH, sequential window acquisition of all theoretical mass spectra; LC-MS/MS, liquid chromatography coupled to tandem mass spectrometry.

### Human plasma proteome dynamics during hyperacute HIV-1 infection

To increase the number of detected proteins, each sample was analysed both neat and after depletion of the 14 most abundant proteins (Fig. 1, Supplementary Information)^29^. In total, 1549 protein profiles were detected, whereof 213 were excluded because they were missing from more than 80% of the samples (Extended Data Fig. 1). Among the remaining 1336 proteins, 379 were collected from neat, and 957 from depleted plasma sample types. Of these, 1028 proteins had unique UniProt IDs and canonical protein form, of which 427 proteins had previously been classified as secreted proteins, actively released into blood plasma, and 601 proteins as intracellular or tissue leakage proteins originating from tissues or dying cells^30,31^. To determine the longitudinal within-host dynamics across V0, V1, and V2 for each of the 1336 identified plasma proteins, kmeans and hierarchical clustering were used. The analysis suggested six different longitudinal expression profiles (Fig. 2a). Of these, two demonstrated a significant decline during hAHI with a rebound to pre-infection levels after hAHI (referred to as “Rapid decrease-rapid increase”, and “Slight decrease-rapid increase”); two demonstrated a decreased levels during and after hAHI (“Gradual decrease”, and “Rapid decrease-slight increase”); one demonstrated an increase during hAHI followed with a decline to pre-infection levels after hAHI (“Rapid increase-rapid decrease”); and one demonstrated a sustained increase during and after hAHI (“Persistent increase”). The general population-based protein dynamics based on mean protein intensities were also determined, indicating that 616 (64%) of the depleted proteins decreased at V1, and that these proteins were mainly involved in cell structure, motility, and transport (Extended Data Fig. 2).

**Fig. 2.**
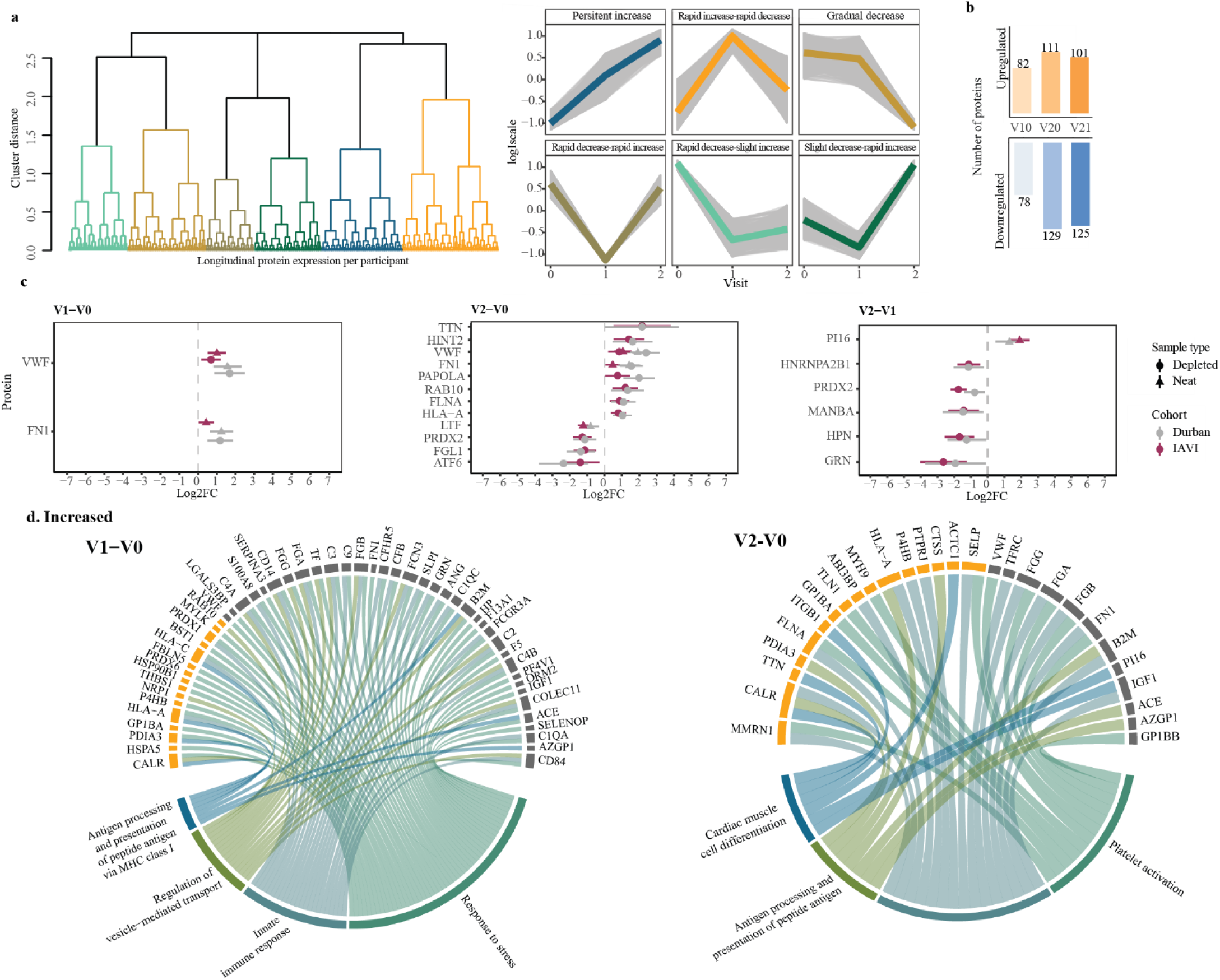
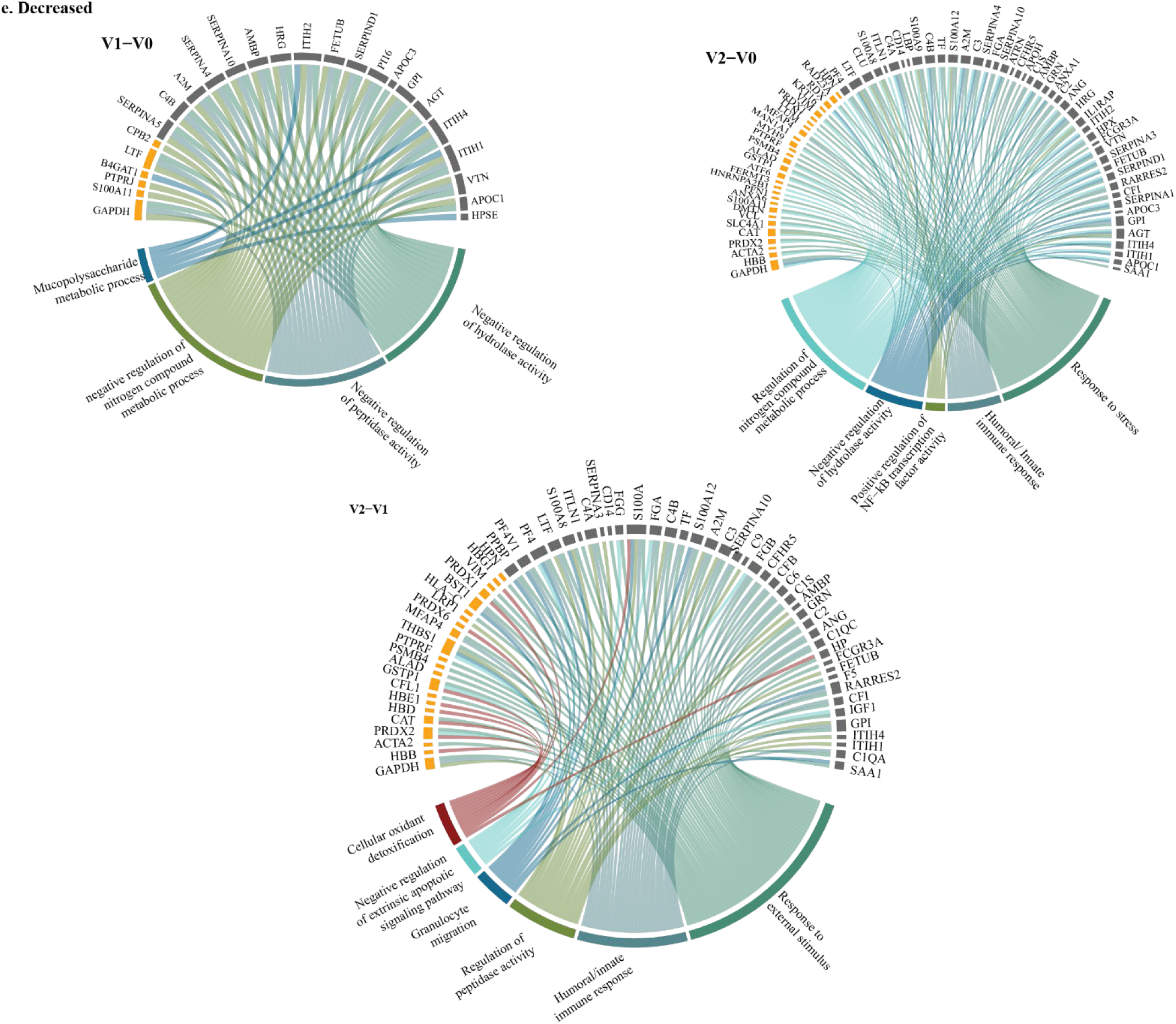
Acute HIV-1 infection alters the human plasma proteome. **a,** Longitudinal protein expression profiles were investigated during hAHI using a dendrogram constructed through hierarchical clustering with complete linkage. A comprehensive analysis of 83,643 protein combination values from all three time points was conducted across 54 study participants, resulting in a total of 1336 profiles. To identify the optimal clusters representing the longitudinal expression profiles for different groups, the elbow method was employed, leading to identification of six distinct clusters. These clusters were color-coded and plotted, with the x-axis denoting the visit number and the y-axis representing the scaled log-intensity per patient. **b,** Bar plot illustrating the comparison in the number of differentially expressed proteins across visit differences. The height of each bar corresponds to the number of proteins while the bar colour varies depending on the visit difference. **c,** Forest plots depicting differentially expressed proteins at two weeks, one-month post-infection, while also accounting for pre-infection levels (V1-V0 and V2-V0, respectively), as well as the difference between visit 2 and visit 1. The plot displays results from linear mixed model analysis, with visit number as a categorical variable (logI ∼ visit + (1|patientid)). The horizontal line represents the confidence interval for the effect size of each protein significantly differentially expressed in both cohorts (depicted in grey for the Durban cohort and maroon for the IAVI cohort). The circle and triangle denote the overall log2FC/effect size for depleted or neat plasma, respectively. Proteins with three or more horizontal lines indicate that those proteins were detected in both depleted and neat plasma (p<0.005; q<0.05). Upregulated proteins in all comparisons are shown on the right side of plot, separated by a vertical dotted grey line, while downregulated proteins are on the left side. **d,e,** circos plots visualising the differentially expressed proteins from different visit differences in a circular layout. The lower ring represents the GO-biological processes associated with the proteins belong to, with each process color-coded for easy identification. The upper rings depict specific classification of these proteins, with proteins secreted in blood shown in orange and the tissue leakage proteins shown in grey.

Next, the proteins that were significantly differentially expressed between the study visits at the individual level were determined. Explorative principal component analysis indicated differential protein expression by cohort and visit, with South African participants clustering separately from East African participants (Extended Data Fig. 3). When adjusting for this potential confounder, 160, 240 and 226 proteins were significantly differentially expressed between visits V1-V0, V2-V0 and V2-V1, respectively (Fig. 2b). Of these, two (V1-V0), 12 (V2-V0), and six (V2-V1) proteins were particularly altered between visits as defined by a log2 fold change (Log2FC) >1 (Fig. 2c; Extended Data Table 2). Of the 160 differentially expressed proteins at V1-V0, 82 proteins were upregulated, of which 57 were overrepresented in gene ontology biological process (GO-BP) terms, including innate immune responses, stress responses, antigen processing or presentation, and regulation of vesicle-mediated transport (Fig. 2d). Of the 78 downregulated proteins, 28 were associated with GO-BP terms such as negative regulation of hydrolase/peptidase activity and cellular metabolic processes (Fig. 2e). Moreover, based on a previous tissue-specific transcriptional signature dataset, significant tissue damage signatures associated with whole blood, oesophagus mucosa and heart were observed in V1 compared to V0 (Extended Data Fig. 4)^32^. When V2 was compared with V0, 111 proteins were upregulated, whereof 40 were linked with different GO-BP terms involved in coagulation cascade/platelet activation, antigen presentation, positive regulation of cell adhesion, and cardiac muscle cell differentiation (Fig. 2d). Of the 129 downregulated proteins, 82 were associated with GO-BP terms and related to hydrolase/peptidase activity, and cellular metabolic processes. Finally, 206 of the 226 proteins that showed differential expression between V1 and V2 could be associated with GO-BP terms, whereof 94 were upregulated. However, none of those were overrepresented in any GO-BP term. In contrast, 67 of the 112 downregulated proteins with GO-BP terms were overrepresented and could be related to immune response, response to external biotic stimulus, complement activation, and regulation of peptidase activity (Fig. 2e). Further analysis indicated that upregulated V2-V1 proteins were associated with tissue damage signatures in adipose, cervical, muscle skeletal and lungs, whereas the downregulated proteins were associated with tissue damage in skin and whole blood tissue (Extended Data Fig. 4). Overall, these findings indicate that HIV-1 infection not only affects inflammatory or immune markers, but also alters protein expression related to different metabolic processes, cell mobility, and apoptosis.

### Decreased levels of Zyxin, Secretoglobin family 1A member 1, and Pro-platelet basic protein during hyperacute HIV-1 infection is associated with ARS

Data on AHI symptoms in the South African cohort were missing. Hence, associations between protein dynamics and ARS were only conducted for participants from the East African cohort (n=33, Fig. 3a). ARS was determined by latent class analysis based on the 11 symptoms including fever, headache, myalgia, fatigue, anorexia, pharyngitis, diarrhoea, night sweats, skin rash, lymphadenopathy, and oral ulcers, as previously described^9^. The analysis suggested that 20 of the 33 participants (61%) had ARS. Participants with ARS had significantly higher prevalence in nine of the eleven symptoms than those without ARS (p<0.05; Fisher exact test, Fig. 3b). Next, Partial Least-Squares Discriminant Analysis (PLS-DA) was used to identify proteins associated with ARS. This analysis demonstrated an average accuracy of 78% (assessed through cross-validation) in predicting ARS in V1-V0 and V2-V0 differences (Fig. 3c). The PLS-DA analysis suggested that 20 differentially expressed proteins with variance importance score above 2 could be potential indicators of ARS (Fig. 3d-e). Notably, approximately half of these proteins at V1-V0 have been shown to be actively secreted into plasma, and are primarily involved in regulation of inflammatory responses, immunity, and host-virus interactions. Proteins identified at V2-V0 were annotated as tissue leakage proteins and are primarily involved in cell motility and signalling (Extended Data Table 3). Next, to confirm the association between the identified proteins and ARS, a linear regression model with age as covariate was applied. The model showed that five of the seven V1-V0 proteins that were determined as potential indicators of ARS in the PLS-DA analysis were significantly associated with ARS (Fig. 3e-f). The linear regression model also indicated that HRG was associated with ARS at V1-V0 (Fig. 3f). A hierarchical clustering analysis of the proteins identified in the PLS-DA and linear regression model analyses was used to identify the top proteins that distinguished participants with and without ARS (Fig. 3g). The analysis indicated clear differential expression of Secretoglobin family 1A member 1 (SCGB1A1), Low affinity immunoglobulin gamma Fc region receptor III-A (FCGR3A), Zyxin (ZYX), and Pro-platelet basic protein (PPBP) between these two groups (Fig. 3g-h). More specifically, significant increases in ZYX, SCGB1A1, and PPBP at V1-V0 were associated with the absence of ARS (Log2FC >±1.5, p<0.005, Fig. 3h). Moreover, the longitudinal expression profiles of these proteins indicated that >70% of individuals with ARS had reduced levels of ZYX, SCGB1, and PPBP at V1-V0 (clusters 1-4, Fig. 3i). ZYX is a cytoskeleton protein that has been shown to be a modulator of inflammatory response in endothelial cells while SCGB1A1 is a pulmonary surfactant anti-inflammatory protein that mediates inflammation and inflammation in the lungs following respiratory distress^33,34^. PPBP, also known as neutrophil-activating peptide-2 or CXCL7, is a cytokine produced by platelets when bound to neutrophil cathepsin G in sites where aggregation of neutrophiles and platelets occur^35^. Furthermore, according to the HIV-1 interactome databases, ZYX and three other proteins have interactions with various HIV-1 proteins that can either enhance or inhibit HIV-1 replication (Supplementary doc 1)^36^. Collectively, these findings highlight a connection between the magnitude of inflammation, innate immunity, and cell motility with the manifestation of ARS.

**Fig. 3.**
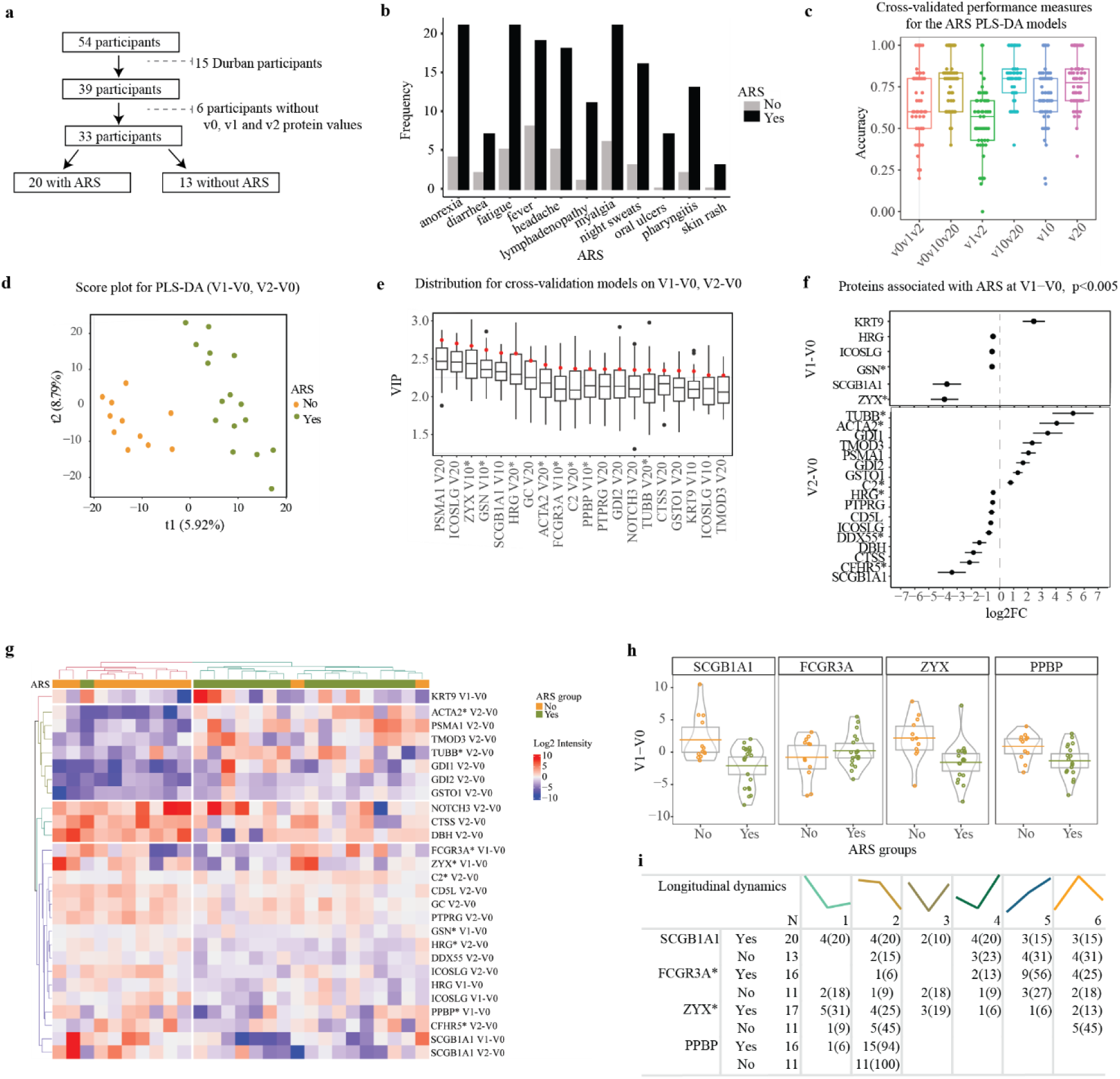
Decreased levels of proteins ZYX, SCGB1 and PPBP two weeks post HIV-1 infection is associated with having ARS. **a,** Flow chart representing the total number of samples used in ARS classification and the exclusion criteria. **b,** Bar graph comparing the distribution of AHI symptoms between volunteers that were defined to be with and without ARS (N=33). ARS was defined based on 11 AHI symptoms, and unobserved linkages between symptoms using Latent Class Analysis. Incremental latent group models were assessed to predict the goodness of fit. The model with two latent groups was the best fit, with the lowest BIC value (660.5) compared to three (678.6), four (699.2), or five (714.7) groups. Study participants were grouped based on their predicted posterior probabilities into those with ARS (N=20/33 (60%)) and those without ARS (13/33 (40%)). **c,** Box plot displaying results of the cross-validated performance measure (accuracy) for the ARS PLS-DA models. These models were trained to predict ARS “Yes” or “No” and evaluated in 10 5-fold cross-validations. For each test set, the performance measures accuracy was computed. Models were constructed based on the following datasets: V0 + V1 + V2; V0 + V1-V0 + V2-V0; V1 + V2; V1-V0 + V2-V0; V1-V0; and V2-V0. **d,** Score plot based on the V1-V0 + V2-V0 dataset (with the highest accuracy value) from (c), indicating the group membership of each sample. There was clear discrimination between the ARS-No (orange) and the ARS-Yes (green) samples on the first (x-axis) and second components (y-axis). Axis labels indicate the percentage of variation explained per component. **e,** Boxplot showing the variable importance in projection (VIP) scores in the PLS-DA model based on V1-V0 for each protein. VIP score summarizes the contribution a variable (protein) makes to the model. This plot identifies the most important proteins for the classification of ARS Yes or No. Proteins with high VIP are more important in providing class separation. Black points represent the full model and the boxplot show distribution for 10 cross-validation models. **f,** Forest plot displaying the proteins associated with ARS using linear mixed models. **g**, Heatmap of proteins associated with ARS based on hierarchical clustering of the V1-V0 and V2-V0 expression of the selected proteins. **h**, Pirate plots showing the V1-V0 protein expression for the top proteins between those with and without ARS. **i**, Table representing the longitudinal protein expression profiles for the top proteins associated with ARS. For each profile and protein, the number (%) of patients with or without ARS were recorded. Abbreviations: ARS, Acute retroviral syndrome; PLS-DA, Partial Least Squares Discriminant analysis; V, visit; V1-V0, difference between visit V1 and V0; V2-V0, difference between visit V2 and V0; V2-V1, difference between visit V2 and V1; Log2FC, log 2-fold change. The asterisk (*) appended to the end of certain protein names indicates proteins detected in neat plasma, while proteins without an asterisk were identified in depleted plasma samples.

### Rho GTPase activating protein 18, Annexin A1, and Lipopolysaccharide binding protein are associated with HIV-1 control

The period during which the VL was monitored varied between study participants. The median follow-up time was four years after the estimated date of infection (EDI). To identify plasma proteins associated with HIV-1 control, 9 study participants who either initiated antiretroviral treatment (ART) within the first year of infection or were followed for less than one year were excluded from the analysis (Fig 4a). The remaining 45 study participants typically showed high levels of viremia following HIV-1 infection, which then subsequently decreased and reached a viral load setpoint after approximately 50 days post the estimated date of infection (EDI, Fig. 4b). The median peak VL was 6.0 (IQR 5.2-6.4) log10 copies/ml at a median of 19 (IQR 15-31) days post EDI, and the median nadir VL was 4.5 log10 copies/ml at a median of 64 (IQR 43-73) days post EDI. Hierarchical clustering was used to group the 45 study participants into two clusters based on their VL profiles during the first year of infection (Fig. 4c). The clusters were defined as viral controllers (VL generally below 10,000 copies/ml for 12 months without ART, n=15), and non-controllers (VL generally above 10,000 copies/ml for 12 months without ART, n=30, Fig. 4d). No clinical parameters, or ARS were associated with viral control (Fig. 4e, Table S2).

**Fig. 4.**
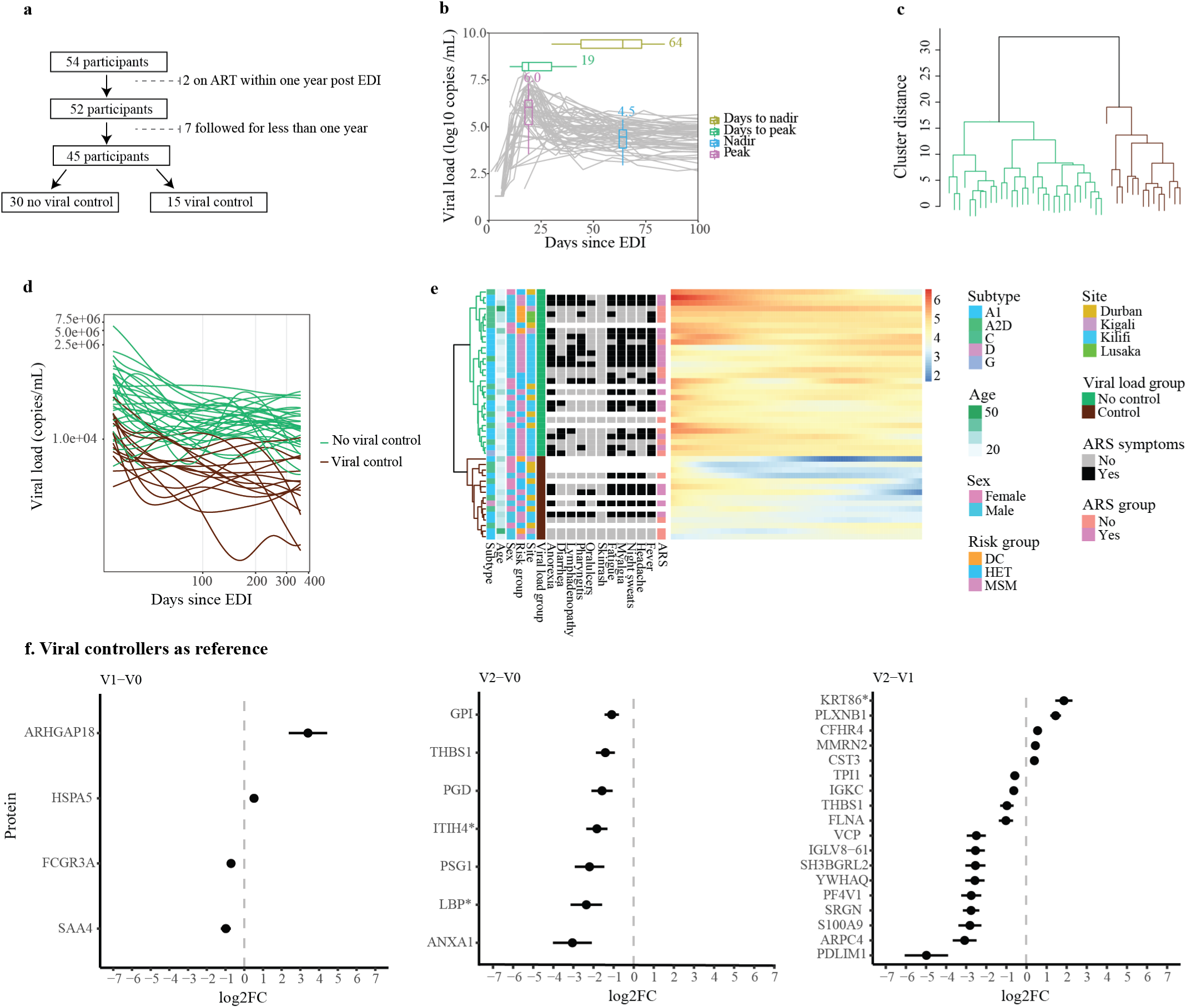
ARHGAP18, ANXA1, and PDLIM1 was associated with HIV-1 control. **a**, Flow chart illustrating the total number of samples used in viral control classification, along with the exclusion criteria. **b**, A plot displaying the viral load measured for all the 54 participants in the first 100 days since EDI. **c**, Dendrogram showcasing complete linkage hierarchical clustering of viral load profiles. Euclidean distances computed from the cubic spline predicted viral load at evenly spread (on transformed scale) time points were used for clustering. The optimal number of clusters was determined using the Silhouette value, and the clustering significance was calculated using multiscale bootstrap resampling. Viral load clusters were based on time 1-12 months (30 to 364 days). Two distinct groups were classified: No viral control (in green) and sustained viral control (in brown). **d**, Plot representing the cubic spline predicted viral load at evenly spread time points. The differentiation between the two viral control groups occurred at a viral load threshold of 10,000 copies/ml. **e**, Heatmap illustrating associations between viral control and various demographic parameters and ARS symptoms. **f**, Forest plot depicitng proteins associated with viral control (clusters based on 1-12 months [complete linkage, Euclidean]) at visit difference V1-V0, V2-V0, and V2-V1. The asterisk (*) appended to certain protein names indicates proteins quantitated from neat plasma, while proteins without an asterisk were identified in depleted plasma samples. Abbreviations: VL, viral load; DC, discordant couple; HET, heterosexual; MSM, men who have sex with men; V, visit. Asterisk (*) indicates proteins quantitated from neat plasma.

Next, linear mixed models (with age and cohort as co-variates) was used to determine the differentially expressed proteins that were associated with viral control. Four, seven and 18 proteins differed significantly between non-viral controllers (cluster 1) and sustained viral controllers (cluster 2) at V1-V0, V2-V0, and V2-V1, respectively (p<0.005; Fig. 4f). The proteins represented acute phase and transport proteins with roles in innate immunity, particularly in the regulation of inflammatory responses and cytokine release (Extended Data Table 4). Specifically, compared with pre-infection levels, Rho GTPase activating protein 18 (ARHGAP18) was significantly higher among viral controllers at V1, whereas Annexin A1 (ANXA1), Lipopolysaccharide binding protein (LBP), and Pregnancy specific beta-1-glycoprotein 1 (PSG1) levels were lower among viral controllers at V2 when compared to the non-viral controllers (Log2FC >2, Fig. 4f). ARHGAP18 activates Rho GTPase, a regulator of cell movement via actin cytoskeleton. Increased levels of ARGHAP18 have been previously coupled to suppression of Rho GTPase thus affecting actin dependent binding of gp120 to the CD4^37^. Annexin is a pro-resolving and anti-inflammatory protein released into blood to neutralise excess protease released by neutrophils following infection^38^. Production of LBP is induced by production of LPS and is a marker of microbial translocation. Increased levels of LBP have been associated to increased activation which could explain why LBP levels are lower among viral controllers^39^. Interestingly, 14 of the proteins associated with viral control have been found to interact with HIV-1 proteins in previous studies, primarily in enhancing virus trafficking to the plasma membrane and infectivity enhancement (supplementary Doc 2)^36^. Collectively, these results indicate a strong relationship between viraemia and cell motility/transport (decreased among those with high viraemia) or innate immunity inflammation (increased among those with high viraemia).

### Hepsin, Protein kinase C beta, and Glutathione S-transferase mu 2 are associated with increased risk of disease progression

Disease progression was defined using CD4+ T-cell responses. Participants contributed a median of 13 (IQR: 10-18) CD4+ T-cell count observations during the 12 months of follow-up after EDI (median CD4+ T-cell count 520, IQR: 404-657 cells/mm^3^). Fast progressors were defined as individuals that reached a CD4+ T-cell count <500 cells/mm^3^ within 12 months from EDI (excluding measurements within the first six weeks), whereas slow progressors were defined as individuals who maintained CD4+ T-cell counts >500 during the same period. Among the 54 participants, 12 (22%) were classified as slow progressors, and 42 (78%) as fast progressors (Extended Data Fig. 5a). Study participants from South Africa had a higher likelihood of slower disease progression compared with East African study participants (p=0.08, Log-rank test, Extended Data Fig. 5b). In a Cox regression model, controlling for age, sex, cohort, and subtype; seven, six, and 14 proteins were associated with faster HIV-1 disease progression at V1-V0, V2-V0, and V2-V1, respectively (p<0.005, Fig. 5c, Extended Data Table 5). Specifically, increased levels of PRKCB (hazard ratio (HR) 1.3, CI=1.1-1.6), HPN (HR=1.4, CI=1.1-1.7), CRHBP (HR=1.2, CI=1.1-1.3), PSMB6 (HR=1.3, CI=1.1-1.5), TXNDC5 (HR=1.2, CI=1.1-1.4), and APOC4 (HR=1.3, CI=1.0-1.2) at V1-V0 were associated with an increased risk of HIV-1 disease progression. Decreased levels of GSTM2 at V1-V0 was associated with a faster disease progression (HR=0.9, CI=0.8-1.0). Interestingly, all these proteins have been shown to interact with the HIV-1 envelope according to the HIV-1 interaction database (supplementary Doc 3)^36^. Moreover, increased levels of ITGB3 (HR=1.3, CI=1.1-1.5), HSPA8 (HR=1.1, CI=1.0-1.2), DDTL (HR=1.2, CI=1.1-1.4) and UBB (HR=1.1, CI=1.0-1.2) at V2-V0 were associated with an increased risk of disease progression. Decreased levels of CD84 (HR=0.9, CI=0.8-0.9) and LTBP1(HR=0.7, CI=0.6-0.9) was associated with decreased risk of disease progression. Notably, all these proteins have been shown to interact with different HIV-1 proteins to mediate cytokine degradation (supplementary Doc 3).

**Fig. 5.**
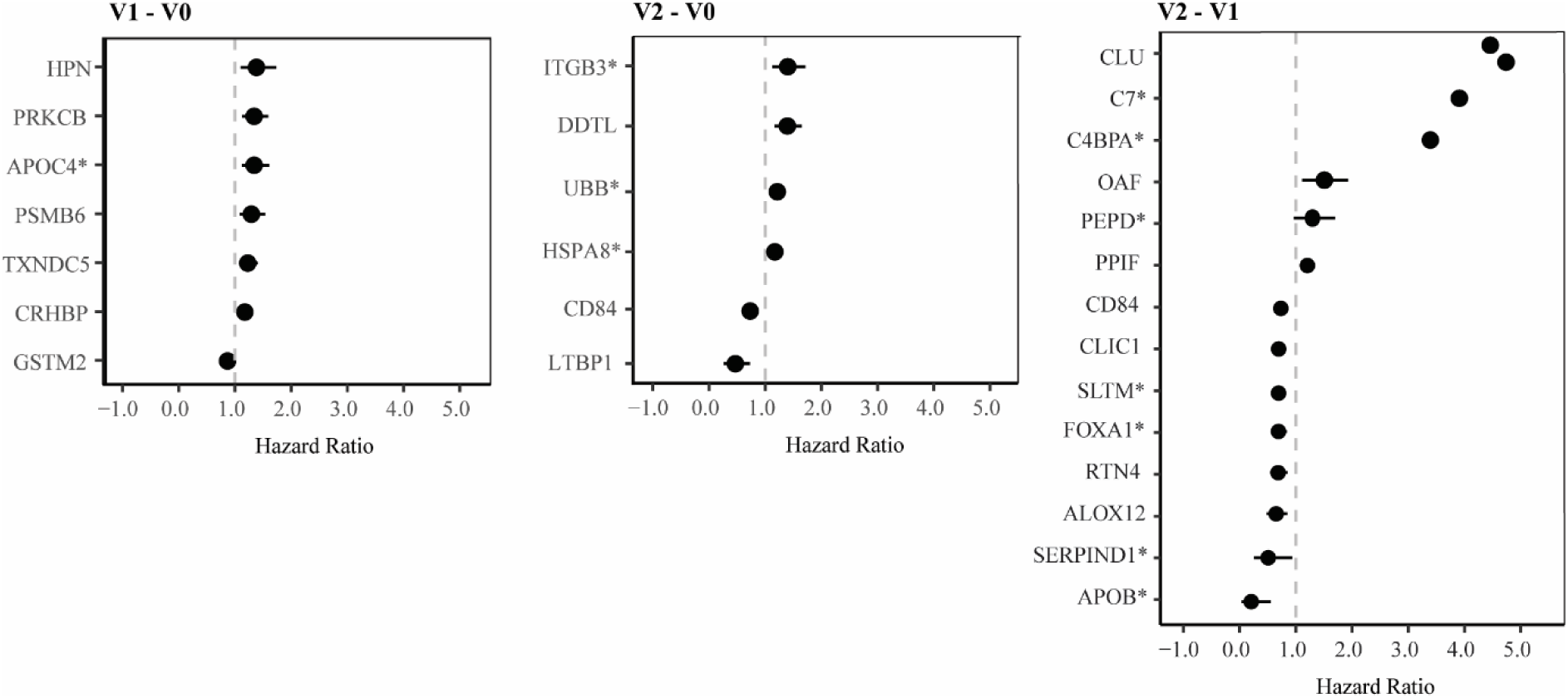
HPN, PRKCB and ITGB3 are associated with increased risk of disease progression. **a**, A forest plot illustrating proteins associated with the time to CD4+ T-cell counts <500 cells/µl, with their corresponding hazard ratios adjusting for age, site of collection, sex and HIV-1 subtype. Protein expressions are based on visit differences V1-V0, V2-V0, and V2-V1 with p-values <0.005, Cox regression models. Asterisk (*) indicates proteins quantitated from neat plasma.

### Longitudinal dynamics of key proteins during hyperacute HIV-1 infection

Finally, a sliding window approach was used to generate spline curves reflecting the population-based average dynamics of identified key proteins in blood plasma during hAHI (Fig. 6). The protein levels were plotted relative to pre-infection levels for each study participants at the day of sample collection post EDI. Key proteins associated with ARS, viral control, and disease progression were selected based on the above analyses. For clarity, proteins with similar dynamics in the group comparisons were excluded from this analysis. The analysis showed an exceptional variability in the trajectory of the different proteins over time and indicated that both the dynamics of overexpressed proteins (defined as *stormers*) and underexpressed proteins (defined as *slumpers*) were common during hAHI.

**Fig. 6.**
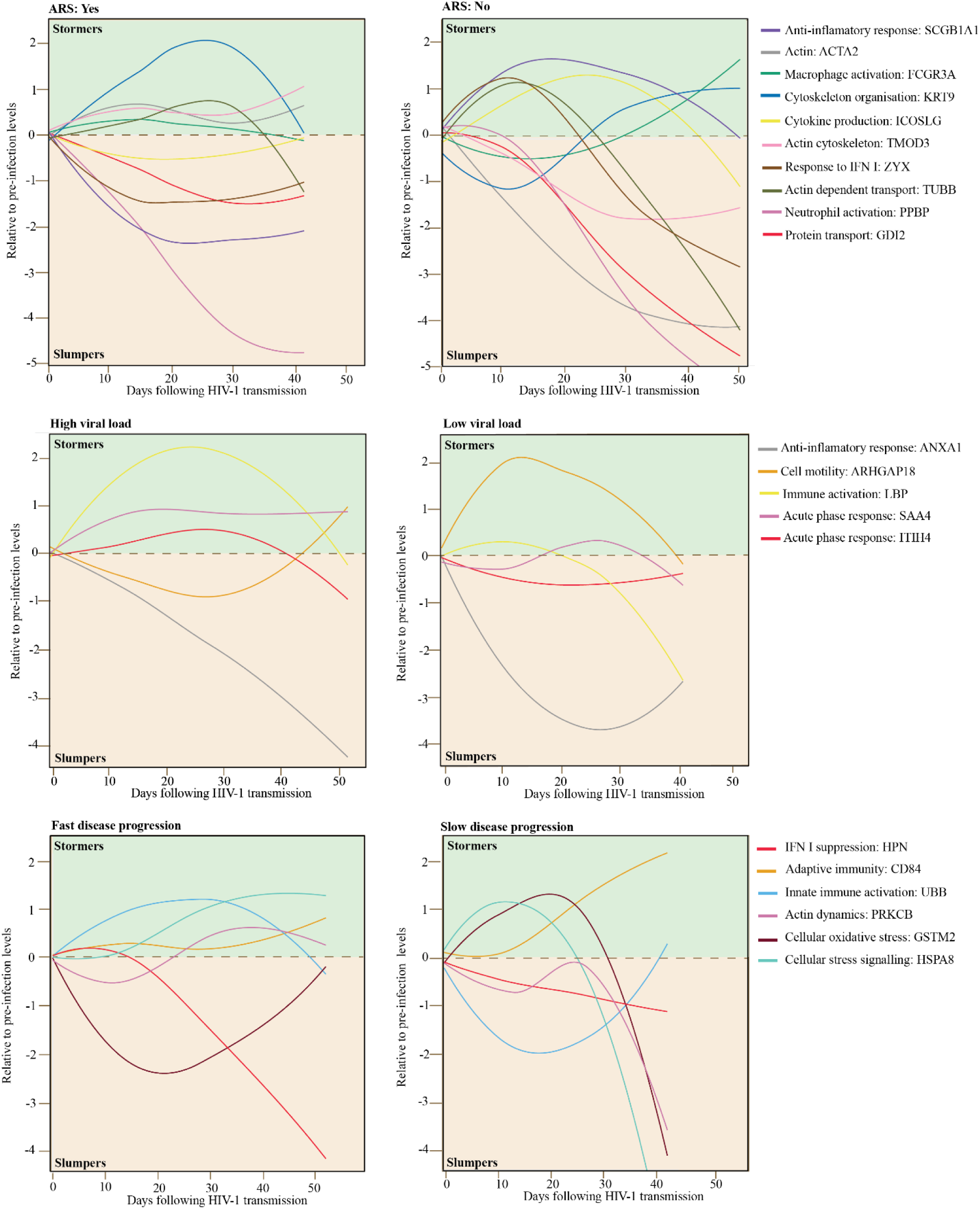
Longitudinal dynamics during hyperacute HIV-1 infection of key differentially expressed proteins associated with ARS, viral load and disease progression. This schematic illustrates the temporal changes in expression for proteins associated with ARS, viral load, and disease progression during acute phase of HIV-1 infection. Protein expression levels were assessed both before infection, and within the two to six weeks following infection in the 54 study participants. Participants were categorized into subgroups based on various outcomes, including ARS (presence or absence), viral control status (controllers or non-controllers), and the rate of disease progression (fast or slow). The x-axis of the graph represents the time in days following infection when plasma samples were collected, while the y-axis represents the mean protein expression levels relative to the pre-infection baseline. Key proteins associated with ARS, viral control and disease progression are depicted using smoothed lines generated through local regression plotting with a span of 1.5. The selected proteins represented the top proteins associated with ARS, viral load, and disease progression To enhance clarity and highlight distinctive patterns, proteins with similar dynamics in the group comparisons were excluded.

## DISCUSSION

Our study represents the largest and most comprehensive longitudinal MS-based study of hAHI. In addition, determination of pre-infection protein levels for each study participant enabled a unique assessment of the protein dynamics relative to HIV-1 uninfected levels at the individual level. The analyses showed significant alterations of the plasma proteome during hAHI, and most changes were transient as the infection progressed. Specifically, elevated expression of SCGB1A1, ZYX, and PPBP during hAHI were associated with absence of ARS; increased levels of ARHGAP18 during hAHI, and decreased levels of ANXA1 and LBP after hAHI were associated with lower viremia; and increased levels of HPN and PRKCB during hAHI, and ITGB3 and DDTL after hAHI, were associated with faster disease progression. These findings are particularly interesting since they represent host factors altered during hAHI that have not been linked to ARS and HIV-1 pathogenesis previously. Previous longitudinal studies in hAHI have focused on the dynamics of pro-inflammatory and antiviral cytokines and chemokines, particularly those immune markers that exhibit increased levels and thereby part of the well-described cytokine storm^9,20,21^. Strikingly, our large-scale analysis of the blood plasma dynamics in hAHI indicated that the differentially expressed proteins associated with ARS, VL, and disease progression was overexpressed (here defined as *stormers*, in analogy with the cytokine storm) to a similar extent as being underexpressed (here defined as *slumpers*). This warrants further studies and it is possible that potential biomarkers and treatment targets may be identified also among slumpers, as among stormers (which have been the main focus so far). Moreover, our study expands beyond studies of acute phase and inflammatory proteins and includes proteins involved in antigen presentation, cell transport, proteolysis, and cytoskeleton modulation during hAHI. For example, vWF and FN1 were the most significantly elevated proteins during hAHI (at peak viremia) and were both persistently elevated after hAHI. vWF reflects persistent endothelial cell activation due to activation of the inflammation/coagulation pathway, whereas FN1 is involved in cell adhesion, cell motility, opsonization, wound healing, and maintenance of cell shape^40^. FN1 also binds to HIV-1 gp120, which has been shown to enhance complement interaction and infection of primary CD4+ T-cells^41^. Moreover, HIV-1 has been shown to modulate the host cytoskeleton dynamics^42^. For example, TTN, a protein that has been implicated in HIV-1 Gag subcellular trafficking, showed increased levels one month after infection, whereas FGL1 levels, a marker of T-cell activation/exhaustion, decreased at the onset of peak viremia, coinciding with the depletion of CD4+ T-cells 3-4 weeks post-infection^43,44^. Furthermore, HNRNPA2B1 levels at four weeks after infection has been associated with virus setpoint levels, and previous studies have suggested that HNRNPA2B1 can interact with viral components, have a critical role in regulating the viral life cycle, and function as a viral DNA sensor initiating IFN production^45,46^.

We have previously shown that a stronger innate immune response in general, and an IP-10 activation in particular, is associated with the manifestation of ARS^9^. In the current study, decreasing levels of ZYX, SCGB1A1, and PPBP during hAHI were associated with ARS. It is possible that decreased levels of SCGB1A1, a pulmonary surfactant protein that influences alveolar macrophage-mediated inflammation, may be linked to lung inflammation and epithelial integrity in individuals with ARS^34,47–49^. Moreover, the associations between SCGB1A1 and ZYX expression and ARS supports previous suggestions of these proteins’ involvement in regulating innate immune responses triggered by viruses^50,51^. Specifically, ZYX binds to the mitochondrial antiviral signalling protein (MAVS) following recognition of cytoplasmic double-stranded RNA by Retinoic acid-inducible gene I (RIG-I)-like receptors. This latter interaction triggers the induction of type I interferon (IFN) expression, which constitute the predominant immune response during hAHI.

Previous studies have suggested that certain cytokines, such as interleukin (IL)-15, IL-7, IL-12p40, IL-12p70, and IFN-gamma, can predict 66% of the variation in viral set-point 12 months after infection^52^. In this study, viral controllers had increased levels of ARHGAP18 during hAHI. Interestingly, ARHGAP18 acts by converting Rho-type GTPases to an inactive GDP-bound state^37^. Rho GTPases play a crucial role in regulating actin cytoskeleton, and HIV-1 and other viruses can induce specific Rho GTPase signalling to facilitate coreceptor binding and transport of virus particles to sites of viral uptake by actin rearrangements^53^. It is therefore plausible that ARHGAP18, which suppresses F-actin polymerization by inhibiting Rho, plays an important role in viral infectivity and fusion, and that reduced actin-dependent binding of HIV-1 to CD4+ T-cells contributes to decreased viral load. Additionally, the levels of the immune inflammation modulator ANXA1 was lower compared with pre-infection levels among viral controllers compared with non-controllers. Indeed, ANXA1 has previously been suggested as a potential therapeutic target towards gut immune dysfunction in HIV-1 infection as a regulator of intestinal mucosal inflammation^54,55^. Furthermore, a gradual increase of ANXA1 expression in simian immunodeficiency virus (SIV) infection has been suggested to facilitate disease progression by impairing inflammatory responses in peripheral blood^54^.

We also found that PRKCB was associated with a faster CD4+ T-cell decline. PRKCB stimulates Nuclear Factor-kappa-B (NF-κB) activation, which regulates B-cell activation, and binds to the HIV-1 promoter, enhancing viral transcription. PRKCB is also involved in cytoskeletal rearrangements necessary for virus entry, further implicating its importance in HIV-1 replication, and the potential exacerbation of CD4+ T-cell decline. Finally, increased levels of the host protease Hepsin was associated with a faster disease progression. This is in line with previous observations that Hepsin suppresses the induction of type I interferons (one of the major defence mechanisms of the human innate immune system towards virus infections)^56^.

The main strength of this study is the well-characterized samples collected before, during and after hAHI. This enabled us to for the first time holistically assess the characteristics of the plasma proteome dynamics in hyperacute HIV-1 infection at both the population and individual patient level, and relative to pre-infection levels. Still, this study is not without limitations. Platelet contamination during plasma separation has been proposed as a potential bias in proteomic studies^57^. Although consistent protocols were employed between cohort sites, the possibility of human error impacting sample collection and handling cannot be fully excluded. Moreover, it is possible that the depletion process may have inadvertently removed some untargeted proteins. Still, the overall quantification of proteins increased 3-fold by this approach, and importantly, the majority of proteins that were detected in both the neat and depleted approaches showed similar results. In summary, this study provides new insights into plasma protein dynamics associated with the complex virus-host interactions and responses that take place when HIV-1 establishes infection in the human host and highlights a similar role of both protein *stormers* and *slumpers* during hAHI. Several novel predictive and prognostic biomarkers associated with ARS, VL responses, and disease progression were identified. The potential implications of these findings are substantial and paves the way for future investigations of diagnostic and prognostic utilities of these biomarkers. Furthermore, our study underscores the need for continued basic research of HIV-1 infection, and other virus infections, to identify biomarkers with potential in early diagnosis, novel and improved treatment strategies of viruses.

## Data Availability

All data produced in the present work are contained in the manuscript

## METHODS

### Study subjects and ethical considerations

Data and samples for this study were obtained from sub-Saharan African (sSA) adults (≥ 18 years old) enrolled in acute and early HIV-1 infection studies. East African volunteers were enrolled in Kenya, Rwanda, and Zambia between 2006 and 2011 under IAVI’s protocols B and C, while South African volunteers were enrolled in Durban between 2007 and 2014 under the FRESH (Females Rising through Education Support and Health) and HIV Pathogenesis Programme (HPP) acute infection cohorts^1–4^. Baseline variables such as date of birth, sex, HIV-1 RNA, HIV-1 p24 antigen, and antibody test results, date of HIV-1 diagnosis, transmission risk group, antiretroviral treatment start date, CD4+ and CD8+ T-cell dynamics, and AHI symptoms were collected. All study participants provided written informed consent for the use of their samples for biomedical research, and all sites received approvals from respective country-specific ethics review boards^1–4^. Additionally, all data and samples were de-identified and anonymised to protect the privacy of the volunteers.

Eligibility for the study included hyperacute HIV-1 infection (hAHI), which was defined as HIV-1 antibody negative and RNA positive (Fiebig stage I) or p24 antigen-positive (Fiebig stage II), corresponding to the period from onset of plasma viremia to peak viral load^5–8^. Matched longitudinal plasma samples were collected at three different visits: (i) visit 0, which was between 22 and 120 days before the estimated date of infection, EDI), (ii) visit 1 which was between 10-14 days post EDI; and (iii) visit 2 which was between 15-42 days post EDI. EDI was defined either as the midpoint between the date of the last negative and first positive HIV antibody test, or 14 days before the date of the first positive p24 antigen test (with a negative antibody test), or 10 days before the date of the first PCR-positive test (with a negative antibody or p24 antigen detection).

### Sample preparation for LC-MS/MS analysis

Blood plasma samples archived at −80°C were obtained for liquid chromatography with tandem mass spectrometry (LC-MS/MS). Both neat and depleted plasma were analysed to increase the number of detected proteins. Depletion refers to removal of the top 14 highly abundant proteins before protein quantification. Details of the specific plasma preparation, LC-MS/MS run conditions, instrumentation and spectral library generation have been documented in supplementary methods.

### DIA/SWATH-MS Targeted Data Extraction

DIA data files were analysed using Spectronaut 15.1 (Biognosys, Schlieren, Switzerland), against the spectral library using the BGS factory default settings. The identifications were filtered at a false discovery rate (FDR) of 1% at both peptide and protein levels. Spectronaut used retention time prediction based on iRT, the m/z dimension in the SWATH-MS data, mass accuracy, and isotopic distribution of fragment ions to identify a peptide. All available transitions were extracted for each targeted peptide, together with their corresponding decoy-transition groups, which were generated by pseudo-reversing the sequence of the targeted peptides.

### Protein quantification and data pre-processing

To derive protein abundances, peptide precursor and fragment ion intensities were used, and the MaxLFQ algorithm (implemented in *iq* R Package) was employed. This algorithm combined multiple peptide ratios to derive optimal protein ratios between pairs of samples, ensuring accuracy and reliability of the data^9,10^. The global distribution of the protein signals was assessed to identify poor quality or low-intensity data. Protein with >80% missing values across samples were removed, and factors such as time of sample collection, date of infection, and date of MS data acquisition were assessed to identify any correlations with missingness. Missing values were imputed by replacing them with a randomly chosen value between one and the minimum global raw intensity value of the protein. The data matrix was then analysed by NormalyzerDE to identify the normalisation method with the least variance in the data, and the normalization was performed based on the results obtained from NormalyzerDE^11^.

### Statistical Analysis

#### Differential expression analysis

To identify differentially expressed proteins (DEPs, proteins that changed over time), a compressed and visualized protein data using principal component analysis (PCA) for several variables including site of collection, HIV-1 subtype, age, and visit number was used. In the following differential analyses across different groups, adjustments were made for principal components (PCs) 1 and 2. Next, the association of each protein with visit number using a linear mixed model with a random intercept for each study participant was tested. A global analysis of variance (ANOVA) test was then conducted to identify the proteins that changed between visits, and *post hoc* tests were performed to determine when the changes occurred. For each protein, global p-values were obtained by likelihood ratio tests of the full model with the effect in question against the model without the effect. Multiple testing correction using the Benjamini-Hochberg’s FDR method with a significance threshold of 5% FDR was applied. A fixed p-value cut-off of p<0.005 for all tests was also applied. The upregulated and downregulated DEPs at different visits were filtered using p<0.005 and plotted in Volcano or Forest plots with 95% CI for log2 fold change.

#### Enrichment or pathway analysis

Protein classifications and annotations were based on subcellular location annotations from the uniProt database, which denotes location and the topology of the mature protein in the cell, and the human protein atlas (HPA) resource^12,13^. To determine whether a set of differentially expressed proteins and their gene ontology (GO) biological processes were statistically different between two biological states (activated and suppressed), enrichment analysis was conducted using Clusterprofiler^14^. The statistical significance of the over-representation was determined based the Fisher’s exact test and Bonferroni’s FDR (p<0.05). The protein list from both neat and depleted plasma was used as the custom background. The top ten enriched terms, along with their respective p-values, were determined.

#### Tissue damage analysis

To evaluate tissue-specific protein expression, a previously established dataset of tissue-enriched transcriptional signatures derived from the Genotype-Tissue Expression (GTEx) project was utilized^15^. The GTEx read counts were transformed into trimmed values and normalized to z-scores for each gene across tissues. Genes with a z-score exceeding three were classified as tissue-enriched. Subsequently, this list of tissue-enriched proteins was employed as the reference database for conducting enrichment analysis with all quantified proteins using Clusterprofiler.

#### Acute Retroviral Syndrome (ARS)

Symptoms during acute HIV infection (AHI) were not part of the study protocol for the FRESH and HPP cohorts and were therefore only recorded for IAVI volunteers using a standardized questionnaire 2-6 weeks after the estimated date of infection^16^. ARS was defined based on 11 symptoms, including fever, headache, myalgia, fatigue, anorexia, pharyngitis, diarrhoea, night sweats, skin rash, lymphadenopathy, oral ulcers. Previous studies have used different definitions for ARS, such as reporting any symptom, ≥2 symptoms, ≥3 symptoms, or a combination of fever with other symptom(s)^17^. However, discrete classification methods may not account for unobserved linkages between symptoms. To address this limitation, latent class analysis (LCA), a structural equation modelling approach, was used to group volunteers based on the number of AHI symptoms and other unobserved linkages. To determine if the proteomic signature could predict or identify ARS, supervised learning approaches, including Partial Least-Squares Discriminant Analysis (PLS-DA, examining all proteins simultaneously) and linear mixed models (considering each protein separately) were used to analyse the proteomic data from all time points and the delta values (protein value change between time points) for each study participant.

#### Disease progression

Disease progression was measured using two endpoints: viral control and CD4+ T-cell decline. For viral control, viral load measurements were taken on various days for each study participant. To compare the viral load profiles between participants, curve fittings were used. All VL measurements from the estimated date of infection until before the start of antiretroviral treatment (ART) were used. VL measurements were log10-transformed, and an optimal smoothing parameter was calculated using leave-one-out cross-validation. A cubic smoothing spline was then fitted separately for each participant, and the Euclidean distance between VL curves was calculated at evenly distributed time points. Participants with observations at the beginning and end of a given time interval were clustered based on Euclidean distance using hierarchical clustering. The optimal number of clusters was determined using the Silhouette method, and clusters were based on a period of 1-12 months.

For CD4+ T-cell decline, a time-to-event analysis using an absolute CD4+ T-cell count of less than 500 from six weeks after EDI as the event was conducted. In addition, the log-rank test was used to assess differences between covariates (p<0.05 was considered statistically significant). The Cox proportional hazards model, adjusting for age, sex, cohort was used to determine the association between each plasma protein and the risk of disease progression. Follow-up time was censored at the initiation of antiretroviral treatment (ART) or the last observed time point if ART was not initiated. The results was presented as Hazard ratios with 95% confidence intervals (CIs), and Kaplan-Meier time-to-event curves.

## Data availability

Proteomics data supporting findings in this study have been deposited to the ProteomeXchange Consortium via PRoteomics IDEntifications (PRIDE) partner repository with the dataset identifier PXD042850 (https://www.ebi.ac.uk/pride/archive/projects/PXD042850).

## Code availability

All original code has been deposited and can be assessed at: https://github.com/NBISweden/SMS-5800-HIV upon request.

## ACKNOWLEGMENTS

The authors thank IAVI for supporting HIV-1 research studies and capacity building initiatives in Kenya, Rwanda, and Zambia. They are also grateful to staff and volunteers from IAVI’s protocol B and C sites in Africa, without whom this work would not have been possible. They also acknowledge the following people for their contributions, and support: Malin Neptin (Department of Translational Medicine, Lund University, Sweden), Ashfaq Ali (NBIS expert), Jakob Wilforss (Department of Immunotechnology, Lund University, Sweden), Christofer Karlsson (Division of Infection Medicine, Department of Clinical Sciences Lund, Faculty of Medicine, Lund University, Sweden), Hong Yan (Department of Clinical Sciences, BioMS, Lund University, Sweden), and Johan Malmström (Division of Infection Medicine, Department of Clinical Sciences Lund, Faculty of Medicine, Lund University, Sweden).

This work was supported by the generous support of the American people through the United States Agency for International Development (USAID). The contents are the responsibility of the study authors and do not necessarily reflect the views of USAID, the National Institutes of Health (NIH), or the US government. This work was also supported by funding from the Swedish Research Council (grant numbers 2016-01417 and 2020-06262 to J.E) and the Swedish Society for Medical Research (grant number SA-2016 to J.E). The authors are also grateful for the support of the Sub-Saharan African Network for TB/HIV-1 Research Excellence (SANTHE), a DELTAS Africa Initiative (grant number DEL-15–006 to A.S.H). with support by the Wellcome Trust (grant number 107752/Z/15/Z), the UK Foreign, Commonwealth & Development Office, through the Developing Excellence in Leadership, Training and Science in Africa (DELTAS Africa) programme. J.N. was funded by the Swedish Research Council (grant numbers 2016-01417 and 2020-06262) and the Medical Faculty at Lund University. A. S. H. was supported by a training fellowship from the Wellcome Trust (209294/Z/17/Z).

Disclaimer. This report was published with permission from the Kenya Medical Research Institute (KEMRI).

## AUTHOR CONTRIBUTIONS

J.E conceived the study. J.E, and A.S.H supervised the research. J.E, A.S.H, and J.N designed the study. J.N and J.E. coordinated experiments and wrote the manuscript with inputs from all authors. J.N, K.G and A.S.H curated the data. J.N, M.R, T.M, S.K performed the sample preparation, LC-MS/MS analysis, and experiment optimisation. J.N, E.J, E.F, M.H, and F.A wrote and optimised the original code used in the study, and performed the data analysis. J.N, E.J, E.F, M.H, F.A, A.S.H, and J.E interpreted the results. All authors reviewed, and approved the final version of the manuscript. The following authors managed the sites to which participants were attended to and where samples were collected, provided clinical expertise and/or provided clinical data for the participant samples: J.H, A.K, E.K, W.K, M.A.P, P.K, S.A, E.H, T.N, J.G, S.R.J, and E.J.S.

## ADDITIONAL INFORMATION

**Supplementary information** is available for this paper.

**Correspondence and requests for materials** should be addressed to J.E.

**Peer review information**

**Reprints and permissions information** is available at http://www.nature.com/reprints.

---

**Extended Data Table 1.** Characteristics of study participants diagnosed with Hyperacute HIV-1 infection. Abbreviations: HIV-1, human immunodeficiency virus type 1; IQR, interquartile range. Risk group data: DC (serodiscordant couples), HET (heterosexual) and MSM (men who have sex with men). Availability of matched pre-infection samples by days from sampling to the estimated date of infection (EDI).

| **Characteristics** | **Total**<br>**(N = 54)** | **Females**<br>**(N = 20)** | **Males**<br>**(N = 34)** |
| --- | --- | --- | --- |
| **Age in years** |  |  |  |
| Median age (IQR) | 24 (22-27) | 22 (21-24) | 26 (22-31) |
| Below age 25 (%) | 32 (59) | 15 (75) | 17 (50) |
| Above age 25 (%) | 22 (41) | 5 (25) | 17 (50) |
| **Country of collection (Site, %)** |  |  |  |
| Kenya (Kilifi) | 32 (59) | 3 (15) | 29 (85) |
| Rwanda (Kigali) | 4 (7) | 2 (10) | 2 (6) |
| Zambia (Lusaka) | 3 (6) | 0 (0) | 3 (9) |
| South Africa (Durban) | 15 (28) | 15 (75) | 0 (0) |
| **HIV-1 subtype (%)** |  |  |  |
| A1 | 31 (57) | 5 (25) | 26 (76) |
| A2D | 1 (2) | 0 (0) | 1 (3) |
| C | 20 (37) | 15 (75) | 5 (15) |
| D | 1 (2) | 0 (0) | 1 (3) |
| G | 1 (2) | 0 (0) | 1 (3) |
| **Risk group (%)** |  |  |  |
| DC | 7 (13) | 2 (10) | 5 (15) |
| HET | 19 (35) | 18 (90) | 1 (3) |
| MSM | 28 (52) | 0 (0) | 28 (82) |
| **Median time in days from EDI (IQR)** |  |  |  |
| Pre-infection (V0) | 62 (28-106) | 59 (41-92) | 71 (22-113) |
| After-Infection – Fiebig stage I (V1) | 10 (7-14) | 7 (7-10) | 9 (10-14) |
| After-Infection – Fiebig stage II (V2) | 31 (28-37) | 29 (28-31) | 32 (29-39) |

**Extended Data Table 2.** Differentially expressed proteins in pre-infection and hyper acute stages of HIV-1. The table presents a comprehensive description of differentially expressed proteins that were observed during acute stages of HIV-1 infection. Significance was determined using a stringent threshold of p<0.005, q<0.005, and log2FC>1 through Linear mixed models, while also ensuring differences were observed in both cohorts. The Protein ID column displays the UniProtKB/Swiss-Prot entry name for each protein, while the Protein description column provides the recommended full protein name from UniProtKB/Swiss-Prot. The Biological process column captures either biological process keywords from UniProtKB/TrEMBL or protein function information provided by the human protein atlas. Secretome location provides information on the predicted location of the protein based on signal peptide and transmembrane region prediction methods listed in HPA, or alternatively, a description of the subcellular location of the mature protein (including isoform locations if available) as described by UNIPROT. Tissue specificity was provided through extracted information from UNIPROT and HPA on the expression of a gene at the mRNA and protein level in cells or tissues of multicellular organisms.

| Protein | Protein description | Biological process | Secretome location | Tissue specificity |
| --- | --- | --- | --- | --- |
| **V1-V0: Increased at V1** |  |  |  |  |
| VWF | Von Willebrand factor | Blood coagulation, Cell adhesion, Haemostasis | Secreted to blood | Plasma |
| FN1 | Fibronectin 1 | Acute phase, Angiogenesis, Cell adhesion, Cell shape | Secreted to blood | Liver |
| **V2-V0: Increased at V2** |  |  |  |  |
| TTN | Titin | cardiac muscle cell development | Leakage | Cardiac and skeletal muscle |
| HINT2 | Histidine triad nucleotide binding 2 | Apoptosis, Lipid biosynthesis, Lipid metabolism, Steroid biosynthesis | Leakage | Liver, Pancreas |
| VWF | Von Willebrand factor | Blood coagulation, Cell adhesion, Haemostasis | Secreted to blood | Plasma |
| FN1 | Fibronectin 1 | Acute phase, Angiogenesis, Cell adhesion, Cell shape | Secreted to blood | Liver |
| PAPOLA | Poly(A) polymerase alpha | mRNA processing | Leakage | Ubiquitous |
| RAB10 | RAB10, member RAS oncogene family | Transport | Leakage | Hippocampus, , testis |
| FLNA | Filamin A | Cilium biogenesis/degradation, actin cytoskeleton reorganisation | Leakage | Ubiquitous |
| HLA-A | Major histocompatibility complex, class I, A | Adaptive immunity, Host-virus interaction, Innate immunity | Leakage | Ubiquitous |
| **V2-V0: Decreased at V2** |  |  |  |  |
| LTF | Lactotransferrin | Immunity, Ion transport, Osteogenesis, Transcription regulation | Secreted to blood | Plasma, tears, saliva, |
| PRDX2 | Peroxiredoxin 2 | T-cell proliferation | Leakage | Ubiquitous |
| FGL1 | Fibrinogen like 1 | Adaptive immunity, regulation of T-cell activation | Secreted to blood | Liver |
| ATF6 | Activating transcription factor 6 | Transcription regulation, Unfolded response | Leakage | Ubiquitous |
| **V2-V1: Increased at V1** |  |  |  |  |
| PI16 | Peptidase inhibitor 16 | cardiac muscle cell development | Secreted to blood | Prostate, urinary bladder, testis |
| **V2-V1: Decreased at V1** |  |  |  |  |
| HNRNPA2B1 | Heterogeneous nuclear ribonucleo A2/B1 | Host-virus interaction, mRNA processing/splicing/ transport | Leakage | Ubiquitous |
| PRDX2 | Peroxiredoxin 2 | T-cell proliferation | Leakage | Ubiquitous |
| MANBA | Mannosidase beta | glyco catabolic process | Leakage | Ubiquitous |
| HPN | Hepsin | positive regulation by host of viral transcription | Leakage | Liver, kidney |
| GRN | Granulin precursor | astrocyte activation involved in immune response | Secreted to blood | Kidney, |

**Extended Data Table 3.** Proteins associated with acute retroviral syndrome. The table presents a comprehensive description of statistically significant proteins at two weeks post HIV-1 infection vs. pre-infection that are associated with ARS. Significance was determined using a stringent threshold of p<0.005, q<0.005, and log2FC>1 through linear mixed modelling, while also ensuring differences were observed in both cohorts. The Protein ID column displays the UniProtKB/Swiss-Prot entry name for each protein, while the Protein description column provides the recommended full protein name from UniProtKB/Swiss-Prot. The Biological process column captures either biological process keywords from UniProtKB/TrEMBL or protein function information provided by the human protein atlas. Secretome location provides information on the predicted location of the protein based on signal peptide and transmembrane region prediction methods listed in HPA, or alternatively, a description of the subcellular location of the mature protein (including isoform locations if available) as described by UNIPROT. Tissue specificity is provided through extracted information from UNIPROT and HPA on the expression of a gene at the mRNA and protein level in cells or tissues of multicellular organisms.

| Protein | Protein description | Biological process | Secretome location | Tissue specificity |
| --- | --- | --- | --- | --- |
| **V1-V0: Increased among those with ARS** |  |  |  |  |
| KRT9 | Keratin 9 | Epithelial cell differentiation | Leakage | Lymphoid tissue |
| **V1-V0: Decreased among those with ARS** |  |  |  |  |
| SCGB1A1 | Secretoglobin family 1A member 1 | Inflammatory response, negative regulation of IFN-gamma/IL-13/IL-4 production | Secreted in other tissues | Club cells |
| ZYX | Zyxin | Inflammatory response, Cell adhesion, Host-virus interaction, response to IFN-gamma | Leakage | Ubiquitous |
| PPBP | Pro-platelet basic protein | Inflammatory response, Chemotaxis | Secreted to blood | Bone marrow, lymphoid tissue |
| FCGR3A | Fc fragment of IgG receptor IIIa | ADCC, regulation of immune response, epithelial cell differentiation | Secreted to blood | Plasma |
| GSN | Gelsolin | Cellular response to interferon-gamma, viral entry into host cell, Cilium biogenesis | Secreted to blood | Plasma |
| ICOSLG | Inducible T-cell costimulator ligand | Adaptive immunity, B-cell activation, Immunity | Membrane | Ubiquitous |
| **V2-V0: Increased among those with ARS** |  |  |  |  |
| ACTA2 | Actin alpha 2 | Cell motility, structure | Leakage | Smooth muscle |
| GDI2 | GDP dissociation inhibitor 2 | Signal transduction, vesicle mediated transport | Leakage | Brain |
| GSTO1 | Glutathione S-transferase omega 1 | Positive regulation of skeletal muscle contraction by regulation of release of sequestered calcium ion | Leakage | Ubiquitous |
| PSMA1 | Proteasome 20S subunit alpha 1 | Inflammatory response, proteasomal protein catabolic process | Leakage | Ubiquitous |
| TMOD3 | Tropomodulin 3 | actin filament organisation | Leakage | Ubiquitous |
| TUBB | Tubulin beta class I | Cytoskeleton-dependent intracellular transport | Leakage | Spleen, thymus, brain |
| **V2-V0: Decreased among those with ARS** |  |  |  |  |
| C2 | Complement C2 | complement activation, innate immunity | Leakage | Liver |
| CTSS | Cathepsin S | adaptive immune response, antigen processing and presentation | Leakage | Lymphoid tissue, bone marrow |
| HRG | Histidine rich glycoprotein | Fibrinolysis, platelet activation, actin cytoskeleton organization | Secreted to blood | Plasma |
| ICOSLG | Inducible T-cell costimulator ligand | Adaptive immunity, B-cell activation, Immunity | Membrane | Ubiquitous |
| PTPRG | Protein tyrosine phosphatase receptor type G | Negative regulation of epithelial cell migration | Membrane | Ubiquitous |

**Extended Data Table 4.**
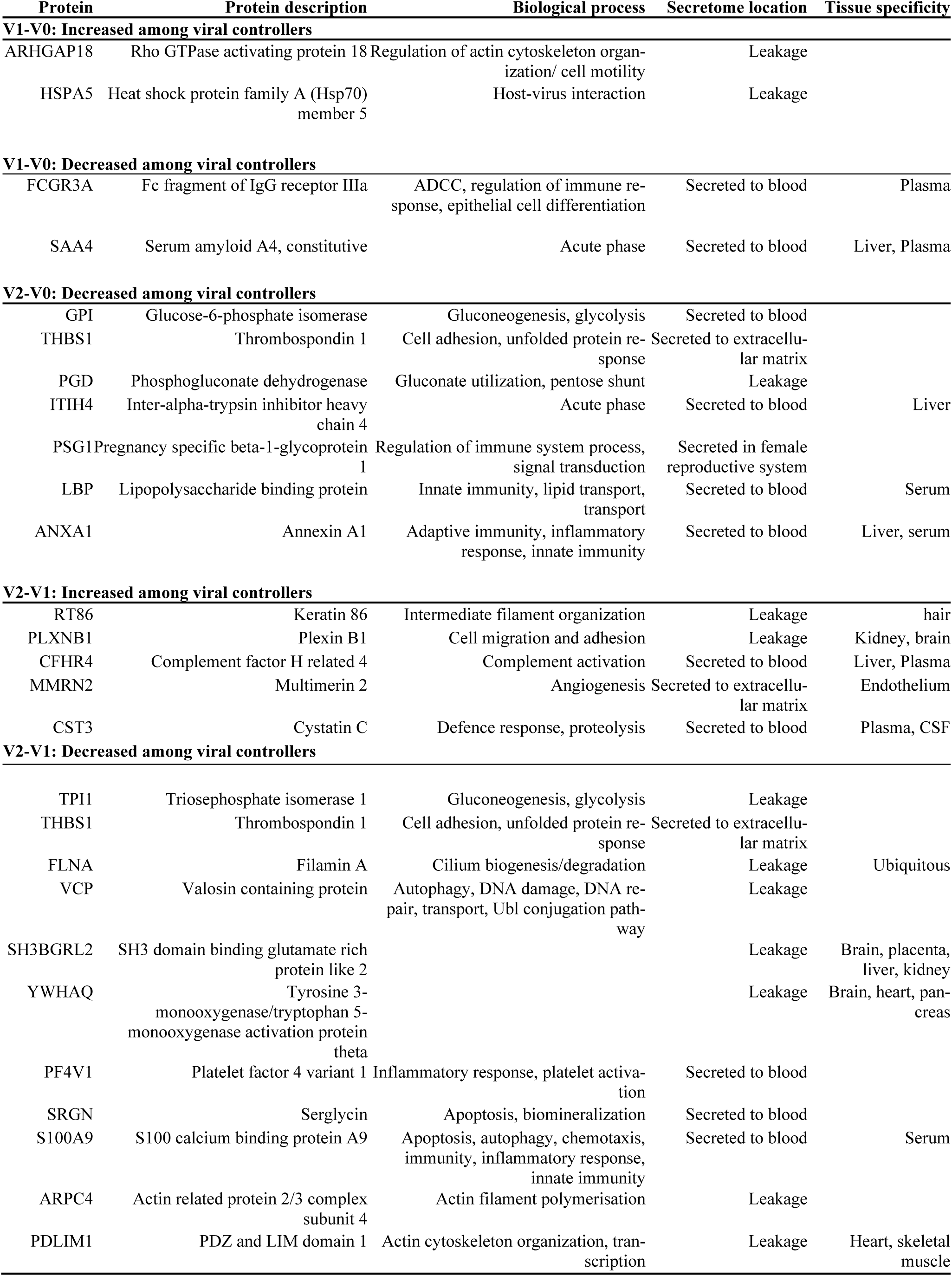
Proteins associated with viral control. The table presents a comprehensive description of statistically significant/ proteins at two weeks post HIV-1 infection vs. pre-infection; one-month post HIV-1 infection vs. pre-infection; one-month vs two-week post HIV-1 infection; that are associated with viral control. To determine significance, a stringent threshold was set at p<0.005, q<0.005, and log2FC>1, while also ensuring differences were observed in both cohorts. The Protein ID column displays the UniProtKB/Swiss-Prot entry name for each protein, while the Protein description column provides the recommended full protein name from UniProtKB/Swiss-Prot. The Biological process column captures either biological process keywords from UniProtKB/TrEMBL or protein function information provided by the human protein atlas. Secretome location provides information on the predicted location of the protein based on signal peptide and transmembrane region prediction methods listed in HPA, or alternatively, a description of the subcellular location of the mature protein (including isoform locations if available) as described by UNIPROT. Tissue specificity is provided through extracted information from UNIPROT and HPA on the expression of a gene at the mRNA and protein level in cells or tissues of multicellular organisms.

**Extended Data Table 5.** Proteins associated with disease progression. The table presents a comprehensive description of statistically significant/ proteins at two weeks post HIV-1 infection vs. pre-infection; one-month post HIV-1 infection vs. pre-infection; one-month vs two-week post HIV-1 infection; that are associated with disease progression. To determine significance, a stringent threshold was at p<0.005, q<0.005, and log2FC>1, while also ensuring differences were observed in both cohorts. The Protein ID column displays the UniProtKB/Swiss-Prot entry name for each protein, while the Protein description column provides the recommended full protein name from UniProtKB/Swiss-Prot. The Biological process column captures either biological process keywords from UniProtKB/TrEMBL or protein function information provided by the human protein atlas. Secretome location provides information on the predicted location of the protein based on signal peptide and transmembrane region prediction methods listed in HPA, or alternatively, a description of the subcellular location of the mature protein (including isoform locations if available) as described by UNIPROT. Tissue specificity is provided through extracted information from UNIPROT and HPA on the expression of a gene at the mRNA and protein level in cells or tissues of multicellular organisms.

| Protein | Protein description | Biological process | Secretome location | Tissue specificity |
| --- | --- | --- | --- | --- |
| **V1-V0: Increase increases risk of progression /Increased among fast progressors** |  |  |  |  |
| HPN | Hepsin | Positive regulation by host of viral transcription | Membrane | Liver, kidney |
| PRKCB | Protein kinase C beta | Adaptive immunity, apoptosis, transcription regulation | Leakage |  |
| APOC4 | Apolipoprotein C4 | Lipid transport, transport | Secreted to blood | Liver, plasma |
| PSMB6 | Proteasome 20S subunit beta 6 | Host-virus interaction | Leakage |  |
| TXNDC5 | Thioredoxin domain containing 5 | Negative regulation of apoptotic process, protein folding | Intracellular and membrane |  |
| CRHBP | Corticotropin releasing hormone binding protein | Inflammatory response | Secreted to blood |  |
| **V1-V0: Decrease increases risk of progression /Decreased among fast progressors** |  |  |  |  |
| GSTM2 | Glutathione S-transferase mu 2 | Regulation of release of sequestered calcium ion | Leakage | Muscle |
| **V2-V0: Increase increases risk of progression /Increased among fast progressors** |  |  |  |  |
| ITGB3 | Integrin subunit beta 3 | Cell adhesion, host-virus interaction | Intracellular and membrane |  |
| DDTL | D-dopachrome tautomerase like |  | Leakage |  |
| UBB | Ubiquitin B | Protein ubiquitination | Leakage |  |
| HSPA8 | Heat shock protein family A (Hsp70) member 8 | Host-virus interaction, mRNA processing, stress response, transcription | Leakage | Ubiquitous |
| **V2-V0: Decrease increases risk of progression /Decreased among fast progressors** |  |  |  |  |
| CD84 | CD84 molecule | Adaptive immunity, autophagy, cell adhesion, innate immunity | Leakage | Hematopoietic tissues |
| LTBP1 | Latent transforming growth factor beta binding protein 1 | Regulation of transforming growth factor beta activation | Secreted to extracellular matrix | Heart |
| **V2-V1: Increase increases risk of progression /Increased among fast progressors** |  |  |  |  |
| CLU | Clusterin | Apoptosis, complement pathway, innate immunity | Secreted to blood | Plasma |
| C7 | Complement C7 | Complement alternate pathway, complement pathway, cytolysis, innate immunity | Secreted to blood | Plasma |
| C4BPA | Complement component 4 binding protein alpha | Complement pathway, innate immunity | Secreted to blood | Plasma |
| **V2-V1: Decrease increases risk of progression /Decreased among fast progressors** |  |  |  |  |
| OAF | Out at first homolog |  | Secreted - unknown location |  |
| PEPD | Peptidase D | Collagen degradation |  |  |
| PPIF | Peptidylprolyl isomerase F | Apoptosis, necrosis | Leakage |  |
| CD84 | CD84 molecule | Adaptive immunity, autophagy, cell adhesion, immunity, innate immunity | Membrane | Hematopoietic tissues |
| CLIC1 | Chloride intracellular channel 1 | Ion transport, transport | Leakage | Heart, placenta, liver, kidney, pancreas |
| SLTM | SAFB like transcription modulator | Apoptosis, transcription, transcription regulation | Leakage |  |
| FOXA1 | Forkhead box A1 | Transcription, transcription regulation | Leakage |  |
| RTN4 | Reticulon 4 | Neurogenesis | Membrane |  |
| ALOX12 | Arachidonate 12-lipoxygenase, 12S type | Fatty acid metabolism, lipid metabolism | Leakage | Vascular smooth muscle cell |
| SERPIND1 | Serpin family D member 1 | Blood coagulation, chemotaxis,<br>haemostasis | Secreted to blood | Liver, plasma |
| APOB | Apolipoprotein B | Cholesterol metabolism, lipid me-<br>tabolism, lipid transport, steroid<br>metabolism, sterol metabolism,<br>transport | Secreted to blood |  |

**Extended Data Fig. 1.**
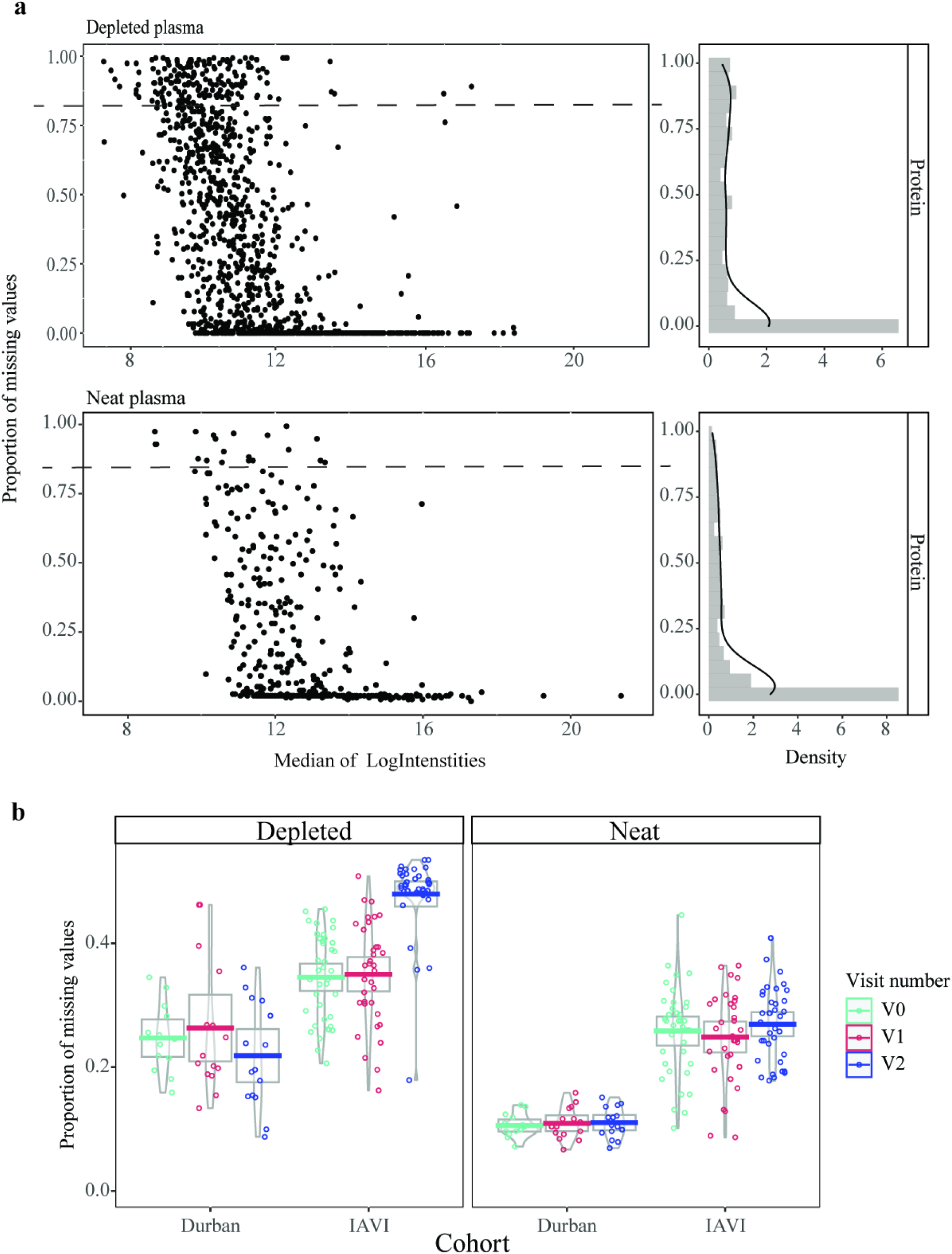
Protein pre-processing and quality control results. **a,** Scatterplot depicitng the protein-wise relationship between median intensities and the proportion of missing values for each sample preparation type in the left panel. Histograms illustrating the frequencies of the proportion are presented alongside with density plots to the right. Protein-wise investigation revealed inverse correlation indicating that proteins with lower median signals generally exhibited more missing values. These trends align with the assumption of missing values are not at random in DIA/SWATH data. **b**, Plot representing the proportion of missing values across cohorts. The IAVI/East African cohort showed higher missingness across all time points when compared to the South African cohort in both depleted and neat plasma samples (p<0.005; Welch Two Sample t-test).

**Extended Data Fig. 2.**
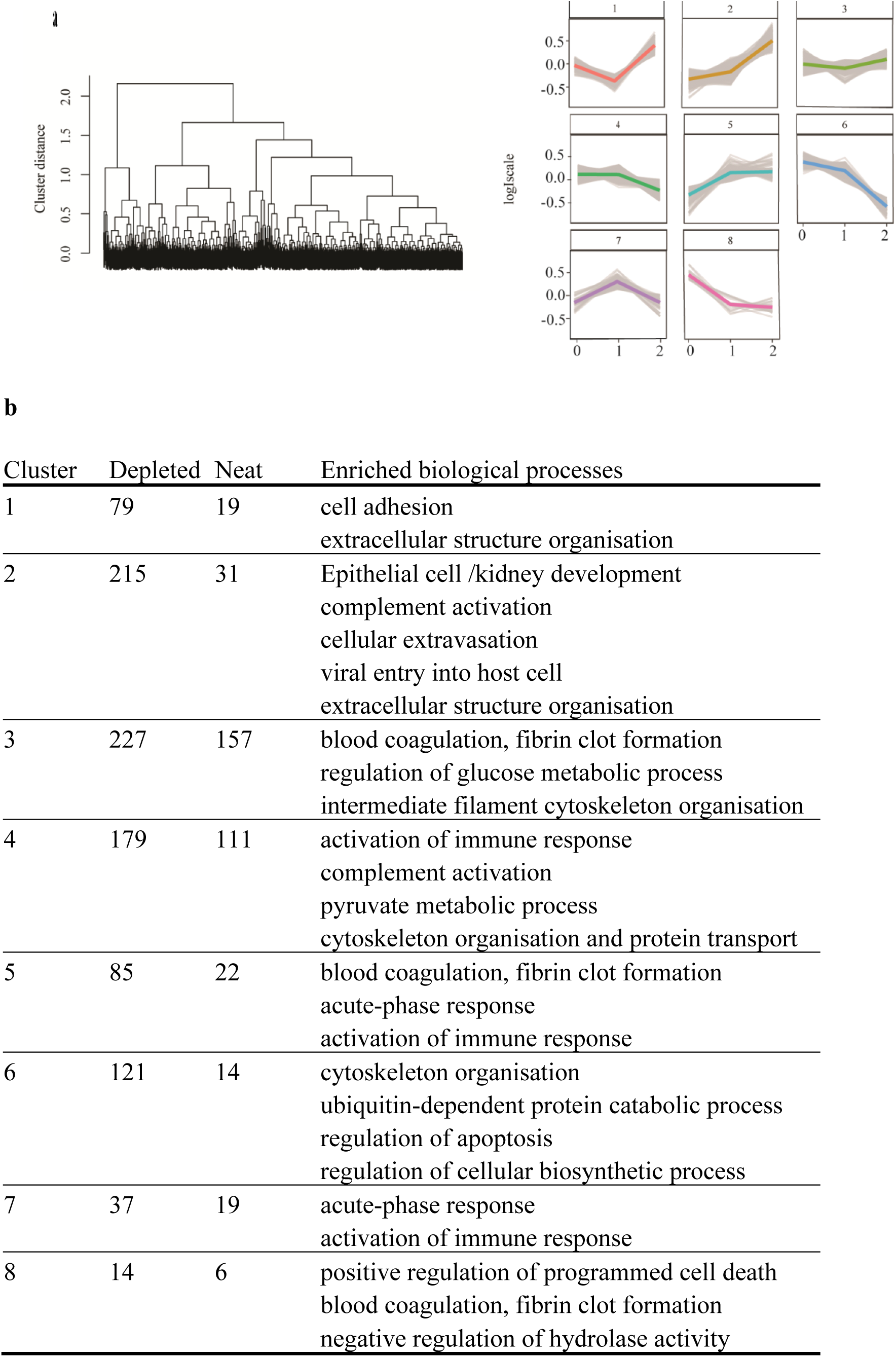
Longitudinal plasma proteome based on mean protein intensities. **a,** Dendrogram illustrating hierarchical clustering with complete linkage for longitudinal protein expression profiles during AHI. The dendrogram was based on the mean protein expression profiles across all 54 patients i.e., 1336 protein combination values from all three time points were analysed. Optimal clusters, indicative of distinct longitudinal expression profiles, were identified using the elbow method, resulting in eight clusters. These clusters were then color-coded and plotted, with the x-axis representing the visit number and the y-axis reflecting the scaled mean log-intensity per protein. **b**, Summary of the number of proteins in both depleted and neat; and enriched biological process terms per cluster.

**Extended Data Fig. 3.**
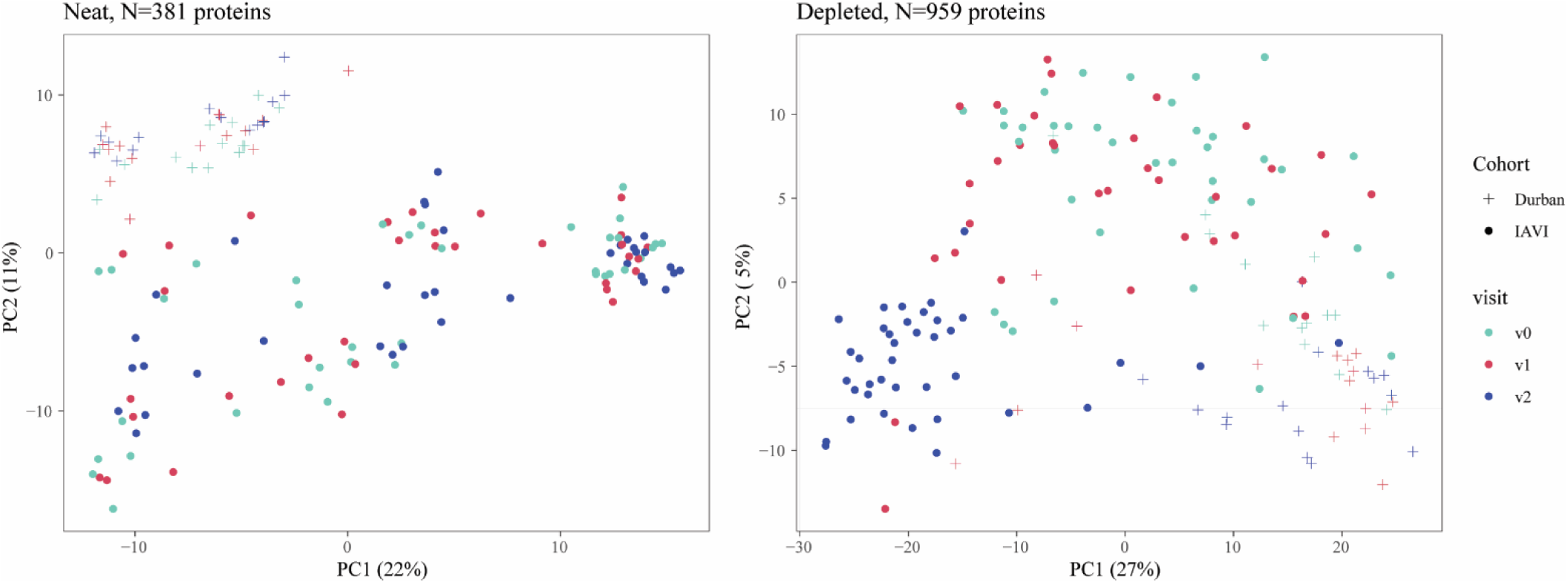
Protein expression different between cohorts. PCA plot illustrating protein expression profiles in AHI samples longitudinally collected from 54 participants across two cohorts. The x- and y-axes represent PCA 1 and 2, respectively, with the explained percentage variance indicated on the axis labels in bracket. Cohorts are distinguished by different symbols: “+” for the Durban/South African cohort and “o” for the IAVI/East African cohort. To preprocess the protein expression data, missing values imputed by replacing each with a randomly chosen value between one and the minimum of the protein that has the missing value. The data was then log-2 transformed and normalised using the Cyclic Loess method (normalizeCyclicLoess function in limma package v3.50.0) as determined by NormalyzerDE.

**Extended Data Fig. 4.**
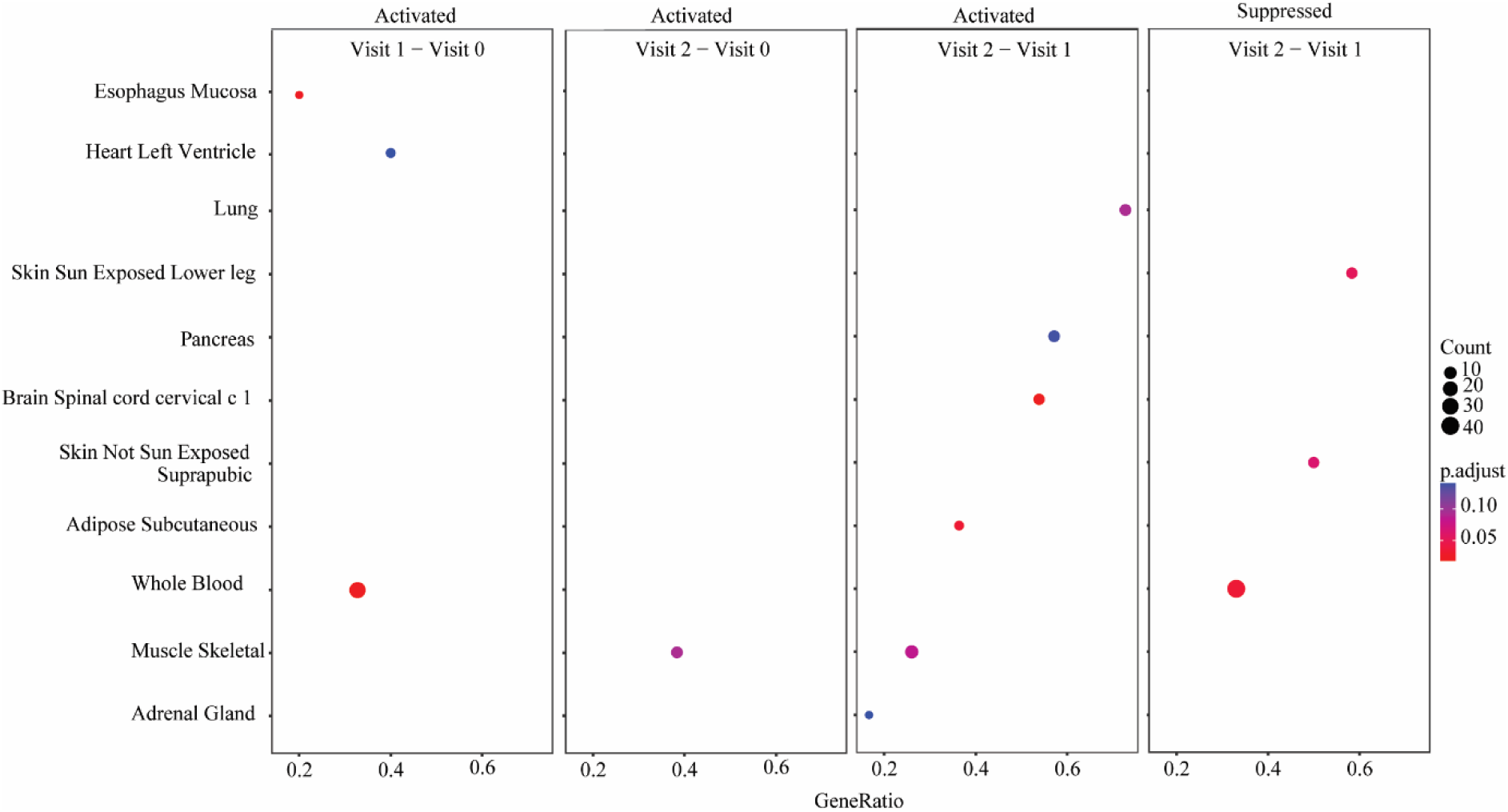
Acute HIV-1 associated tissue damage signatures. Dot plot illustrating the results of gene set enrichment analysis for tissue damage signatures obtained using a tissue damage library. Each dot represents a gene set and is positioned based on its enrichment score and statistical significance. The size of the dot corresponds to the significance of enrichment, with larger dots indicating higher significance. The color of the dot represents the adjusted p-value associated with the protein set. Dots located below the significance threshold (p<0.05) indicate positive enrichment, signifying overrepresentation of the gene set in the analysed data. The tissue damage signatures are displayed on the y-axis, while the x-axis represents the Protein/GeneRatio.

**Extended Data Fig. 5.**
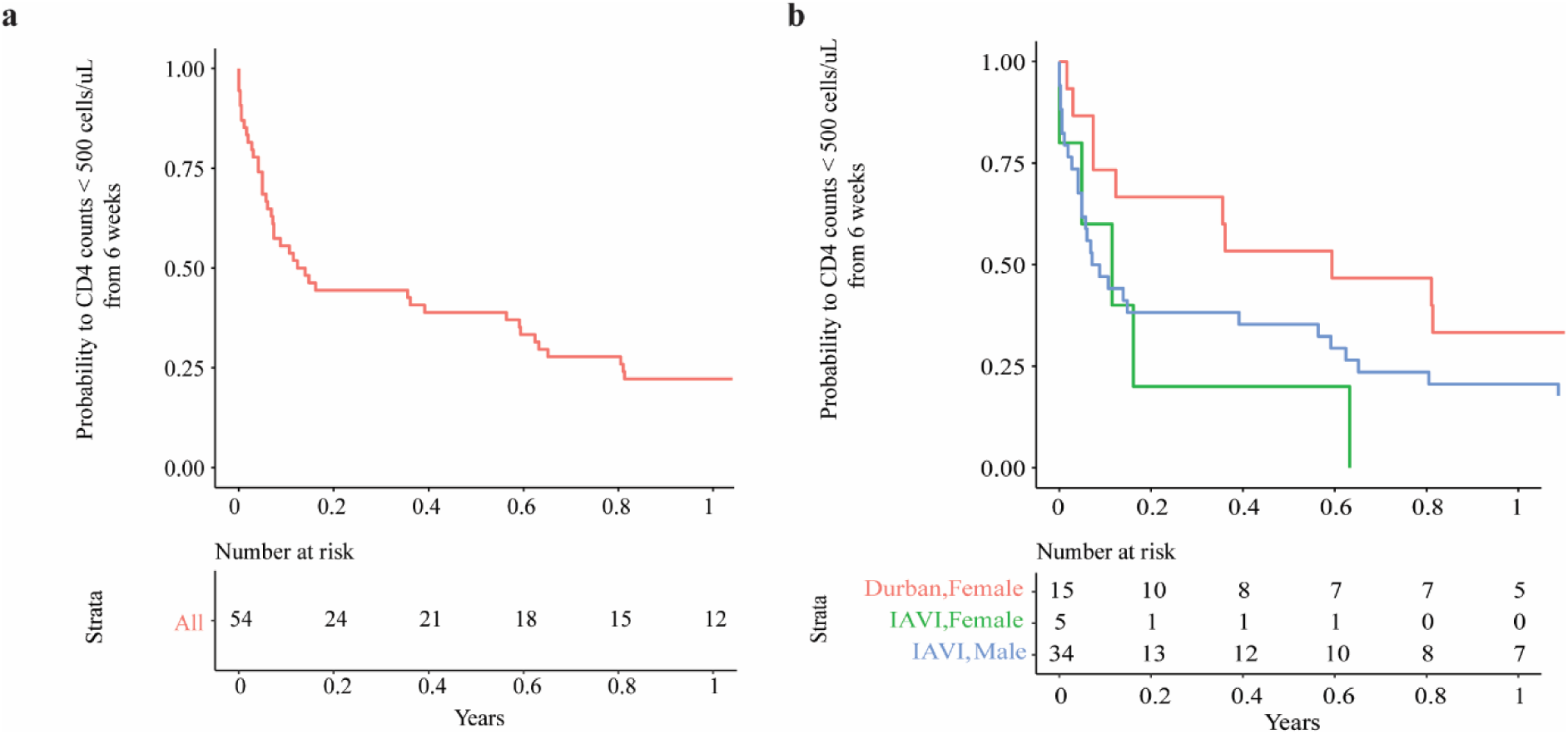
Classification of disease progression. **a**, Kaplan-Meier plot displaying the estimated probability of survival against time to CD4+ T-cell counts <500 cells/µl, starting from six weeks post estimated date of infection to ART start date. The number of study participants at risk at regular time intervals is shown at the bottom of the figure. **b**, A Kaplan-Meier plot displaying the differences in survival between females and male study participants and a p-value of 0.08 from the Log rank test is indicative of no significant difference in survival between males and females.

## REFERENCES

1 Zhong, W. et al. Whole-genome sequence association analysis of blood proteins in a longitudinal wellness cohort. Genome Med 12, 53, doi:10.1186/s13073-020-00755-0 (2020).

2 Captur, G. et al. Plasma proteomic signature predicts who will get persistent symptoms following SARS-CoV-2 infection. eBioMedicine 85, doi:10.1016/j.ebiom.2022.104293 (2022).

3 Palma Medina, L. M., et al. Targeted plasma proteomics reveals signatures discriminating COVID-19 from sepsis with pneumonia. Respiratory Research 24, 62, doi:10.1186/s12931-023-02364-y (2023).

4 Al-Nesf, M. A. Y. et al. Prognostic tools and candidate drugs based on plasma proteomics of patients with severe COVID-19 complications. Nature Communications 13, 946, doi:10.1038/s41467-022-28639-4 (2022).

5 Cohen, M. S., Shaw, G. M., McMichael, A. J. & Haynes, B. F. Acute HIV-1 Infection. New England Journal of Medicine 364, 1943–1954, doi:10.1056/NEJMra1011874 (2011).

6 McMichael, A. J., Borrow, P., Tomaras, G. D., Goonetilleke, N. & Haynes, B. F. The immune response during acute HIV-1 infection: clues for vaccine development. Nature Reviews Immunology 10, 11–23, doi:10.1038/nri2674 (2010).

7 Ndhlovu, Z. M. et al. Magnitude and Kinetics of CD8+ T Cell Activation during Hyperacute HIV Infection Impact Viral Set Point. Immunity 43, 591–604, doi:10.1016/j.immuni.2015.08.012 (2015).

8 Ndhlovu, Z. M. et al. Augmentation of HIV-specific T cell function by immediate treatment of hyperacute HIV-1 infection. Sci Transl Med 11, doi:10.1126/scitranslmed.aau0528 (2019).

9 Hassan, A. S. et al. A Stronger Innate Immune Response During Hyperacute Human Immunodeficiency Virus Type 1 (HIV-1) Infection Is Associated With Acute Retroviral Syndrome. Clinical Infectious Diseases 73, 832–841, doi:10.1093/cid/ciab139 (2021).

10 Robb, M. L., Eller, L. A. & Rolland, M. Acute HIV-1 Infection in Adults in East Africa and Thailand. The New England journal of medicine 375, 1195, doi:10.1056/NEJMc1609157 (2016).

11 Sanders, E. J. et al. Differences in acute retroviral syndrome by HIV-1 subtype in a multicentre cohort study in Africa. AIDS (London, England) 31, 2541–2546, doi:10.1097/qad.0000000000001659 (2017).

12 Tindall, B. et al. Characterization of the acute clinical illness associated with human immunodeficiency virus infection. Arch Intern Med 148, 945–949 (1988).

13 Kazer, S. W., Walker, B. D. & Shalek, A. K. Evolution and Diversity of Immune Responses during Acute HIV Infection. Immunity 53, 908–924, doi:10.1016/j.immuni.2020.10.015 (2020).

14 Brenchley, J. M. et al. CD4+ T cell depletion during all stages of HIV disease occurs predominantly in the gastrointestinal tract. J Exp Med 200, 749–759, doi:10.1084/jem.20040874 (2004).

15 Levesque, M. C. et al. Polyclonal B cell differentiation and loss of gastrointestinal tract germinal centers in the earliest stages of HIV-1 infection. PLoS Med 6, e1000107, doi:10.1371/journal.pmed.1000107 (2009).

16 Lavreys, L. et al. Higher set point plasma viral load and more-severe acute HIV type 1 (HIV-1) illness predict mortality among high-risk HIV-1-infected African women. Clin Infect Dis 42, 1333–1339, doi:10.1086/503258 (2006).

17 Lefrère, J. J. et al. The risk of disease progression is determined during the first year of human immunodeficiency virus type 1 infection. J Infect Dis 177, 1541–1548, doi:10.1086/515308 (1998).

18 Lindbäck, S. et al. Viral dynamics in primary HIV-1 infection. Karolinska Institutet Primary HIV Infection Study Group. AIDS (London, England) 14, 2283–2291, doi:10.1097/00002030-200010200-00009 (2000).

19 Deeks, S. G. et al. Research priorities for an HIV cure: International AIDS Society Global Scientific Strategy 2021. Nature Medicine 27, 2085–2098, doi:10.1038/s41591-021-01590-5 (2021).

20 Muema, D. M. et al. Association between the cytokine storm, immune cell dynamics, and viral replicative capacity in hyperacute HIV infection. BMC Medicine 18, doi:10.1186/s12916-020-01529-6 (2020).

21 Stacey, A. R. et al. Induction of a striking systemic cytokine cascade prior to peak viremia in acute human immunodeficiency virus type 1 infection, in contrast to more modest and delayed responses in acute hepatitis B and C virus infections. J Virol 83, 3719–3733, doi:10.1128/jvi.01844-08 (2009).

22 Deutsch, E. W. et al. Advances and Utility of the Human Plasma Proteome. Journal of Proteome Research 20, 5241–5263, doi:10.1021/acs.jproteome.1c00657 (2021).

23 Uhlén, M. et al. Proteomics. Tissue-based map of the human proteome. Science 347, 1260419, doi:10.1126/science.1260419 (2015).

24 Kamali, A. et al. Creating an African HIV clinical research and prevention trials network: HIV prevalence, incidence and transmission. PloS one 10, e0116100, doi:10.1371/journal.pone.0116100 (2015).

25 Price, M. A. et al. Cohort Profile: IAVI’s HIV epidemiology and early infection cohort studies in Africa to support vaccine discovery. International journal of epidemiology 50, 29–30, doi:10.1093/ije/dyaa100 (2021).

26 Dong, K. L. et al. Detection and treatment of Fiebig stage I HIV-1 infection in young at-risk women in South Africa: a prospective cohort study. The lancet. HIV 5, e35–e44, doi:10.1016/s2352-3018(17)30146-7 (2018).

27 Bassett, I. V. et al. Screening for acute HIV infection in South Africa: finding acute and chronic disease. HIV Med 12, 46–53, doi:10.1111/j.1468-1293.2010.00850.x (2011).

28 Fiebig, E. W. et al. Dynamics of HIV viremia and antibody seroconversion in plasma donors: implications for diagnosis and staging of primary HIV infection. AIDS (London, England) 17, 1871–1879, doi:10.1097/00002030-200309050-00005 (2003).

29 Tu, C. et al. Depletion of Abundant Plasma Proteins and Limitations of Plasma Proteomics. Journal of Proteome Research 9, 4982–4991, doi:10.1021/pr100646w (2010).

30 Consortium, T. U. UniProt: the Universal Protein Knowledgebase in 2023. Nucleic Acids Research 51, D523–D531, doi:10.1093/nar/gkac1052 (2022).

31 Uhlén, M., Fagerberg, L., Hallström, B. M., Lindskog, C. & al., e. Tissue-based map of the human proteome. 347, 1260419, doi:doi:10.1126/science.1260419 (2015).

32 Arthur, L. et al. Cellular and plasma proteomic determinants of COVID-19 and non-COVID-19 pulmonary diseases relative to healthy aging. Nature Aging 1, 535–549, doi:10.1038/s43587-021-00067-x (2021).

33 Wójtowicz, A. et al. Zyxin mediation of stretch-induced gene expression in human endothelial cells. Circ Res 107, 898–902, doi:10.1161/circresaha.110.227850 (2010).

34 Xu, M., Yang, W., Wang, X. & Nayak, D. K. Lung Secretoglobin Scgb1a1 Influences Alveolar Macrophage-Mediated Inflammation and Immunity. Frontiers in Immunology 11, doi:10.3389/fimmu.2020.584310 (2020).

35 Cohen, A. B., Stevens, M. D., Miller, E. J., Atkinson, M. A. & Mullenbach, G. Generation of the neutrophil-activating peptide-2 by cathepsin G and cathepsin G-treated human platelets. Am J Physiol 263, L249–256, doi:10.1152/ajplung.1992.263.2.L249 (1992).

36 NIH. HIV-1 Human Interaction Database, <https://www.ncbi.nlm.nih.gov/genome/viruses/retroviruses/hiv-1/interactions/> (2023).

37 Maeda, M. et al. ARHGAP18, a GTPase-activating protein for RhoA, controls cell shape, spreading, and motility. Mol Biol Cell 22, 3840–3852, doi:10.1091/mbc.E11-04-0364 (2011).

38 Vago, J. P. et al. Proresolving Actions of Synthetic and Natural Protease Inhibitors Are Mediated by Annexin A1. The Journal of Immunology 196, 1922–1932, doi:10.4049/jimmunol.1500886 (2016).

39 Younas, M. et al. Microbial Translocation Is Linked to a Specific Immune Activation Profile in HIV-1-Infected Adults With Suppressed Viremia. Frontiers in Immunology 10, doi:10.3389/fimmu.2019.02185 (2019).

40 Schapkaitz, E., Libhaber, E., Jacobson, B. F., Meiring, M. & Büller, H. R. von Willebrand factor propeptide-to-antigen ratio in HIV-infected pregnancy: Evidence of endothelial activation. Journal of Thrombosis and Haemostasis 19, 3168–3176, 10.1111/jth.15502 (2021).

41 Greco, G., Pal, S., Pasqualini, R. & Schnapp, L. M. Matrix Fibronectin Increases HIV Stability and Infectivity1. The Journal of Immunology 168, 5722–5729, doi:10.4049/jimmunol.168.11.5722 (2002).

42 Naghavi, M. H. & Goff, S. P. Retroviral proteins that interact with the host cell cytoskeleton. Curr Opin Immunol 19, 402–407, doi:10.1016/j.coi.2007.07.003 (2007).

43 Cooper, J. et al. Filamin A Protein Interacts with Human Immunodeficiency Virus Type 1 Gag Protein and Contributes to Productive Particle Assembly *<sup></sup>. Journal of Biological Chemistry 286, 28498–28510, doi:10.1074/jbc.M111.239053 (2011).

44 Wang, J. et al. Fibrinogen-like Protein 1 Is a Major Immune Inhibitory Ligand of LAG-3. Cell 176, 334–347.e312, doi:10.1016/j.cell.2018.11.010 (2019).

45 Bhattarai, K. & Holcik, M. Diverse roles of heterogeneous nuclear ribonucleoproteins in viral life cycle. Frontiers in Virology 2, doi:10.3389/fviro.2022.1044652 (2022).

46 Zhang, X., Flavell, R. A. & Li, H.-B. hnRNPA2B1: a nuclear DNA sensor in antiviral immunity. Cell Research 29, 879–880, doi:10.1038/s41422-019-0226-8 (2019).

47 Pang, M. et al. Recombinant CC16 protein inhibits the production of pro-inflammatory cytokines via NF-κB and p38 MAPK pathways in LPS-activated RAW264.7 macrophages. Acta Biochimica et Biophysica Sinica 49, 435–443, doi:10.1093/abbs/gmx020 (2017).

48 Dierynck, I., Bernard, A., Roels, H. & De Ley, M. Potent inhibition of both human interferon-gamma production and biologic activity by the Clara cell protein CC16. Am J Respir Cell Mol Biol 12, 205–210, doi:10.1165/ajrcmb.12.2.7865218 (1995).

49 Xu, M., Yang, W., Wang, X. & Nayak, D. K. Lung Secretoglobin Scgb1a1 Influences Alveolar Macrophage-Mediated Inflammation and Immunity. Frontiers in immunology 11, 584310, doi:10.3389/fimmu.2020.584310 (2020).

50 Huang, S. et al. SOCS Proteins Participate in the Regulation of Innate Immune Response Caused by Viruses. Front Immunol 11, 558341, doi:10.3389/fimmu.2020.558341 (2020).

51 Kouwaki, T. et al. Zyxin stabilizes RIG-I and MAVS interactions and promotes type I interferon response. Sci Rep 7, 11905, doi:10.1038/s41598-017-12224-7 (2017).

52 Roberts, L. et al. Plasma cytokine levels during acute HIV-1 infection predict HIV disease progression. AIDS (London, England) 24, 819–831, doi:10.1097/QAD.0b013e3283367836 (2010).

53 Van den Broeke, C., Jacob, T. & Favoreel, H. W. Rho’ing in and out of cells: viral interactions with Rho GTPase signaling. Small GTPases 5, e28318, doi:10.4161/sgtp.28318 (2014).

54 Sena, A. A. et al. Divergent Annexin A1 expression in periphery and gut is associated with systemic immune activation and impaired gut immune response during SIV infection. Sci Rep 6, 31157, doi:10.1038/srep31157 (2016).

55 Babbin, B. A. et al. Annexin A1 regulates intestinal mucosal injury, inflammation, and repair. J Immunol 181, 5035–5044, doi:10.4049/jimmunol.181.7.5035 (2008).

56 Hsin, F., Hsu, Y. C., Tsai, Y. F., Lin, S. W. & Liu, H. M. The transmembrane serine protease hepsin suppresses type I interferon induction by cleaving STING. Sci Signal 14, doi:10.1126/scisignal.abb4752 (2021).

57 Geyer, P. E. et al. Plasma Proteome Profiling to detect and avoid sample-related biases in biomarker studies. EMBO Molecular Medicine 11, doi:10.15252/emmm.201910427 (2019).

## METHOD REFERENCES

1 Kamali, A. et al. Creating an African HIV clinical research and prevention trials network: HIV prevalence, incidence and transmission. PloS one 10, e0116100, doi:10.1371/journal.pone.0116100 (2015).

2 Price, M. A. et al. Cohort Profile: IAVI’s HIV epidemiology and early infection cohort studies in Africa to support vaccine discovery. International journal of epidemiology 50, 29–30, doi:10.1093/ije/dyaa100 (2021).

3 Dong, K. L. et al. Detection and treatment of Fiebig stage I HIV-1 infection in young at-risk women in South Africa: a prospective cohort study. The lancet. HIV 5, e35–e44, doi:10.1016/s2352-3018(17)30146-7 (2018).

4 Bassett, I. V. et al. Screening for acute HIV infection in South Africa: finding acute and chronic disease. HIV Med 12, 46–53, doi:10.1111/j.1468-1293.2010.00850.x (2011).

9 Cox, J. et al. Accurate proteome-wide label-free quantification by delayed normalization and maximal peptide ratio extraction, termed MaxLFQ. Molecular & cellular proteomics : MCP 13, 2513–2526, doi:10.1074/mcp.M113.031591 (2014).

10 Pham, T. V., Henneman, A. A. & Jimenez, C. R. iq: an R package to estimate relative protein abundances from ion quantification in DIA-MS-based proteomics. Bioinformatics 36, 2611–2613, doi:10.1093/bioinformatics/btz961 (2020).

11 Willforss, J., Chawade, A. & Levander, F. NormalyzerDE: Online Tool for Improved Normalization of Omics Expression Data and High-Sensitivity Differential Expression Analysis. Journal of Proteome Research 18, 732–740, doi:10.1021/acs.jproteome.8b00523 (2019).

12 Consortium, T. U. UniProt: the Universal Protein Knowledgebase in 2023. Nucleic Acids Research 51, D523–D531, doi:10.1093/nar/gkac1052 (2022).

13 Uhlén, M., et al. Proteomics. Tissue-based map of the human proteome. Science 347, 1260419, doi:10.1126/science.1260419 (2015).

14 Wu, T. et al. clusterProfiler 4.0: A universal enrichment tool for interpreting omics data. The Innovation 2, 100141, 10.1016/j.xinn.2021.100141 (2021).

15 Arthur, L. et al. Cellular and plasma proteomic determinants of COVID-19 and non-COVID-19 pulmonary diseases relative to healthy aging. Nature Aging 1, 535–549, doi:10.1038/s43587-021-00067-x (2021).

16 Sanders, E. J. et al. Differences in acute retroviral syndrome by HIV-1 subtype in a multicentre cohort study in Africa. AIDS (London, England) 31, 2541–2546, doi:10.1097/qad.0000000000001659 (2017).

17 Crowell, T. A. et al. Acute Retroviral Syndrome Is Associated With High Viral Burden, CD4 Depletion, and Immune Activation in Systemic and Tissue Compartments. Clin Infect Dis 66, 1540–1549, doi:10.1093/cid/cix1063 (2018).

