## Supplemental Information for "Dynamics of the blood plasma proteome during hyperacute HIV-1 infection"

Affiliations

**SUPPLEMENTARY METHODS**

**Sample preparation for LC-MS/MS analysis**

*Neat plasma*

The protein digestion process was optimized for 1 μl (1 mg/ml) of plasma. First, 50μl of digestion buffer (8 M urea, in 100 mM ammonium bicarbonate) was added to the plasma. Next, proteins were reduced with 5 mM Tris(2-carboxyethyl) phosphine, pH 7.0 for 60 minutes at 37 °C, and then alkylated with 25 mM iodoacetamide (Sigma) at room temperature for 30 minutes in the dark. The mixture was diluted with 100 mM ammonium bicarbonate to achieve a final urea concentration below 1.5 M, and trypsin (1/100, w/w, Sequencing Grade Modified Trypsin, Porcine; Promega) was added for overnight digestion (at least 9 hours) at 37 °C. Digestion was halted using 5% trifluoracetic acid (TFA) (Sigma) to pH 2-3, and the peptides were purified and desalted using SOLAμ^TM^ reverse phase solid phase extraction plates (Thermo Fisher Scientific), following the manufacturer's instructions. After washing with 50% acetonitrile with 0.1% TFA, the solvents were evaporated using a vacuum concentrator (Genevac, miVac), and the peptides were resuspended in 50μl HPLC-water (Fisher Chemical) containing 2% acetonitrile and 0.1% formic acid (Sigma).

An equivalent sample amount of 1μg peptide was then injected into the column, and the samples were spiked with indexed retention time peptides (iRT) peptides before LC-MS analysis. The LC-MS analysis for neat plasma was conducted using an EASY-nLC 1200 ultra-HPLC system coupled to a Q Exactive HF-X mass spectrometer (Thermo Fisher Scientific) with the following column setting: trap column (PepMap100 C18 3 μm; 75 μm × 2 cm; Thermo Fisher Scientific), EASY-Spray column (ES803, column temperature 45 °C; Thermo Fisher Scientific), and linear gradient from 5% to 38% over 90 or 120 minutes at a flow rate of 350 nl/min. The DIA-44 variable windows + 90 min NL gradientnLC was used for LC-MS/MS analysis. The full MS resolution was set to 60,000 at 200 m/z, and the mass range was set to 350–1650 m/z.

For neat plasma, a spectral library was established using the Pulsar search engine integrated into Spectronaut 15.1(Biognosys, Schlieren, Switzerland) with the factory default settings. Briefly, peptides from 24 immunodepleted plasma were pooled and fractionated with off-line high-pH reversed-phase chromatography (Thermo Fisher Scientific). The fractions were then subjected to DDA analysis, and the raw DIA and DDA data were loaded directly into Pulsar and searched against the human reference proteome acquired from Uniprot Homo sapiens Database (December 2019, 42,410 entries). The generated library consisted of 15,896 precursors: 8616 peptides, 781 protein groups, and 1272 proteins.

*Depleted plasma*

To enhance the detection of medium and low range proteins in plasma, a depletion strategy was employed to remove high abundant proteins. Specifically, 95% of the top 14 most abundant proteins, including HSA, albumin, IgG, IgA, IgM, IgD, IgE kappa and lambda light chains, alpha-1-acid glycoprotein, alpha-1-antitrypsin, alpha-2-macroglobulin, apolipoprotein A1, fibrinogen, haptoglobin, and transferrin, were removed to increase the sensitivity of detection for low abundance proteins. The plasma samples were processed using High Select™ Top14 Abundant Protein Depletion Mini Spin Columns according to the manufacturer’s protocol, which involved the use of resins containing highly specific immobilized antibodies. Briefly, four μl plasma was added to the column, mixed for 10 min at room temperature, and the eluent was collected by centrifugation at 1,000 × g for 2 min.

The depleted plasma was then subjected to digestion using the filter-aided sample preparation (FASP) method. To denature the proteins, 8M urea in 100 mM Ammonium bicarbonate was used, and disulfide bridges were reduced with freshly prepared 35mM dithiothreitol (DTT). The proteins were then alkylated with 55mM iodoacetamide (IAA) to allow immediate access of the trypsin to the internal cleavage sites. Ultrafiltration facilitated by centrifugation was employed to remove DTT and other low-molecular-weight components. The filters were washed with 100mM Ammonium bicarbonate in between the different steps to ensure the removal of any remaining DTT or IAA.

Prior to protein digestion, the concentration of the protein was measured using the Qubit™ 4 Fluorometer (#Q33239, Thermo Fisher Scientific, Waltham, MA, USA) and the Invitrogen Qubit™ Protein BR Assay Kit (#Q33211, Thermo Fisher Scientific, Waltham, MA, USA). Sequencing Grade Modified Trypsin (1µg/µl,#V5111, Promega, Madison, WI, USA) was added to 30 µg protein for each sample in 100mM ammonium bicarbonate and incubated at 37°C for 16 hours. The digestion was halted by adding 10% trifluoroacetic acid (TFA) (#302031, Sigma- Aldrich, Merck, Darmstadt, Germany), and the pH was checked using pH strips. The peptide concentration was measured using the Nanodrop™ 2000 spectrophotometer (#ThermoND-2000, Fisher Scientific, Waltham, MA, USA) after digestion.

For LC-MS analysis, an equal amount of 1 µg peptide was injected into the column, and the samples were spiked with indexed retention time peptides (iRT) before analysis. The Dionex Ultimate 3000 RSLCnano UPLC coupled to an Exploris 480 mass spectrometer with FAIMS (Thermo Fischer Scientific) was used for LC-MS analysis of depleted plasma. The column settings were as follows: trap column (PN 164535), anal column (ES802A) LC-MS/MS analysis (DIA): DIA-26 variable windows + FAIMS with 2 CVs (-45V & -60V), 90 min NL gradient. nLC: – non-linear gradient, 1 min at 5% B, in 75 min up to 25% B, in 9 min up to 32% B, in 6 min up to 45% B, in 2 min up to 95% B, 5 min at 95% B and 12 min equilibration at 5% B full MS - resolution: 120.000, normalized AGC target: 300%, maxIT: 45 ms, 380-1100 m/z,DIA – 26 windows with variable width, resolution: 30.000, normalized AGC target: 1000%, maxIT: auto, NCE: 32, centroid.

Due to differences in instrumentation, a separate spectral library was generated for depleted plasma. A total of 10 immunodepleted plasma samples were pooled, fractionated (N=8), and analyzed by DDA (DDA-max speed [1.7s + 1.3s cycle time] FAIMS with two CVs [-45V and -60V], 90 min NL gradient). The generated raw DIA data was combined with the DDA data and searched against Uniprot Homo sapiens Database (July 2020, 42,386 entries) using pulsar. The resulting spectral library generated using Spectronaut 15.1(Biognosys, Schlieren, Switzerland), contained 17,727 precursors: 12,529 peptides, 1,294 protein groups, and 2,259 proteins.

**DIA**/**SWATH-MS Targeted Data Extraction**

DIA data files were analysed using Spectronaut 14.10.201222.47784 (Biognosys, Schlieren, Switzerland), against the spectral library using the BGS factory default settings. The identifications were filtered at an FDR of 1% at both peptide and protein levels. Spectronaut used retention time prediction based on iRT (28), the m/z dimension in the SWATH-MS data, mass accuracy and isotopic distribution of fragment ions to identify a peptide. For each targeted peptide, all available transitions were extracted, along with their corresponding decoy-transition groups, which were generated by pseudo-reversing the sequence of the targeted peptides.

**HIV-1 Subtyping**

HIV-1 env sequences (V1-V3, approximately 940 base pairs) were generated, and the HIV-1 subtype was phylogenetically determined as previously described^1^. Briefly, the general time-reversible (GTR) model of nucleotide substitution with gamma-distributed rate heterogeneity was used to infer maximum likelihood trees. Branch support was assessed using the approximate likelihood ratio test based on the Shimodaira-Hasegawa (aLRT-SH) method, with branch support of ≥ 0.90 considered significant^2^.

**SUPPLEMENTARY TABLES**

**Table S1. Differentially expressed proteins across visit differences.** The table lists the number of proteins that were significantly different in abundance pre-infection (V0), two weeks post infection (V1) and one-month post infection (V2). Using visit difference enabled us to cater for individual variability in protein expression before and after HIV-1 infection.

| Differentially expressed proteins | V1-V0 | V2-V0 | V2 -V1 |
| --- | --- | --- | --- |
| Up – regulated | 82 | 111 | 101 |
| Down – regulated | 78 | 129 | 125 |
| Total | **160** | **240** | **226** |

**Table S2. Association between ARS, clinical parameters, and viral control.** The table displays the results of various tests assessing the association of viral control with several variables, including acute retroviral syndrome (ARS), site, HIV-1 transmission risk group, age, sex, and HIV-1 subtype. Each row represents a specific variable, and the corresponding test was used to evaluate the association between viral controllers and non-viral controllers. Variables investigated include ARS, site, HIV-1 transmission risk group, age, sex, and HIV-1 subtype. The test column lists the several tests used to assess the association between the variable and viral control. The p-value associated with the test, representing the statistical significance of the results and providing evidence of no association, is shown in the last column. The numbers in parentheses indicate the percentage of participants for each variable among viral control groups.

| **Variable** | **Non-viral controllers** | **Viral controllers** | **Test** | **p-value** |
| --- | --- | --- | --- | --- |
| **N** | 30 | 15 | None |  |
| **ARS=Yes (%)** | 14 (58.3) | 4 (50) | Fisher | 0.703 |
| Fever=Yes (%) | 20 (83.3) | 4 (50) | Fisher | 0.152 |
| Headache=Yes (%) | 13 (54.2) | 5 (62.5) | Fisher | 1 |
| Nightsweats=Yes (%) | 13 (54.2) | 5 (62.5) | Fisher | 1 |
| Myalgia=Yes (%) | 16 (66.7) | 5 (62.5) | Fisher | 1 |
| Fatigue=Yes (%) | 17 (70.8) | 5 (62.5) | Fisher | 0.681 |
| Skinrash=Yes (%) | 0 (0) | 1 (12.5) | Fisher | 0.25 |
| Oralulcers=Yes (%) | 4 (16.7) | 2 (25) | Fisher | 0.625 |
| Pharyngitis=Yes (%) | 10 (41.7) | 3 (37.5) | Fisher | 1 |
| Lymphadenopathy=Yes (%) | 9 (37.5) | 1 (12.5) | Fisher | 0.38 |
| Diarrhea=Yes (%) | 8 (33.3) | 1 (12.5) | Fisher | 0.386 |
| Anorexia=Yes (%) | 16 (66.7) | 4 (50) | Fisher | 0.433 |
| **Site (%)** |  |  | Fisher | 0.338 |
| Durban | 6 (20) | 6 (40) |  |  |
| Kigali | 2 (6.7) | 2 (13.3) |  |  |
| Kilifi | 20 (66.7) | 7 (46.7) |  |  |
| Lusaka | 2 (6.7) | 0 (0) |  |  |
| **Risk group (%)** |  |  | Fisher | 0.337 |
| DC | 4 (13.3) | 2 (13.3) |  |  |
| HET | 8 (26.7) | 7 (46.7) |  |  |
| MSM | 18 (60) | 6 (40) |  |  |
| **Sex=Male (%)** | 22 (73.3) | 7 (46.7) | Chi square | 0.152 |
| **Age (mean (SD))** | 25.52 (6.74) | 27.65 (6.81) | Mann-Whitney U | 0.0766 |
| **Subtype (%)** | (%) |  | Fisher | 0.649 |
| A1 | 18 (60) | 8 (53.3) |  |  |
| A2D | 1 (3.3) | 0 (0) |  |  |
| C | 10 (33.3) | 6 (40) |  |  |
| D | 0 (0) | 1 (6.7) |  |  |
| G | 1 (3.3) | 0 (0) |  |  |

**REFERENCES**

1	Esbjornsson, J., Mild, M., Mansson, F., Norrgren, H. & Medstrand, P. HIV-1 Molecular Epidemiology in Guinea-Bissau, West Africa: Origin, Demography and Migrations. *PLoS ONE* **6**, doi:10.1371/journal.pone.0017025 (2011).

2	Guindon, S. *et al.* New Algorithms and Methods to Estimate Maximum-Likelihood Phylogenies: Assessing the Performance of PhyML 3.0. *Systematic biology* **59**, 307-321, doi:10.1093/sysbio/syq010 (2010).
